# Prediction of Mutations and Outcome in Gastrointestinal Stromal Tumors with Deep Learning: A Multicenter, Multinational Study

**DOI:** 10.64898/2026.02.02.26345350

**Authors:** A. Bonetti, V.L. Le, Z. I. Carrero, F. Wolf, M. Gustav, S.W. Lam, L. Vanhersecke, P. Sobczuk, F. Le Loarer, M. Lenarcik, P. Rutkowski, J. M. van Sabben, N. Steeghs, H. van Boven, I. Machado, S. Bagué, S. Navarro, E. Medina-Ceballos, C. Agra, F. Giner, G. Tapia, A. Hernández-Gallego, G. Civantos Jubera, M. Cuatrecasas, S. Lopez-Prades, R.E. Perret, I. Soubeyran, E. Khalifa, L. Blouin, E. Wardelmann, A. Meurgey, P. Collini, A. Voloshin, Y. Yatabe, H. Hirano, A. Gronchi, T. Nishida, O. Bouché, J.F. Emile, C. Ngo, P. Hohenberger, C. Cotarelo, J. Jakob, J.V.M.G. Bovee, H. Gelderblom, A. Szumera-Cieckiewicz, M. Jean-Denis, J. Bollard, N. Lassau, A. Lecesne, J.Y. Blay, A. Italiano, A. Crombé, J.M. Coindre, J. N. Kather

## Abstract

**Background:** Gastrointestinal stromal tumor (GIST) is the most common gastrointestinal mesenchymal tumor, driven by tyrosine-protein kinase KIT and platelet-derived growth factor receptor A (PDGFRA) mutations. Specific variants, such as KIT exon 11 deletions, carry prognostic and therapeutic implications, whereas wild-type (WT) variants derive limited benefit from tyrosine kinase inhibitors (TKIs). Given the limited reproducibility of established clinicopathological risk models, deep learning (DL) applied to whole-slide images (WSIs) emerged as a promising tool for molecular classification and prognostic assessment.

**Patients and methods:** We analyzed 8398 GIST cases from 21 centers in 7 countries, including 7238 with molecular data and 2638 with clinical follow-up. DL models were trained on WSIs to predict mutations, treatment sensitivity, and recurrence-free survival (RFS).

**Results:** DL predicted mutational status in GIST from WSIs, with area under the curve (AUC) of 0.87 for *KIT*, 0.96 for *PDGFRA*. High performance was observed for subtypes, including KIT exon 11 delinss 557–558 (0.67) and *PDGFRA* exon 18 D842V (0.93). For therapeutic categories, performance reached 0.84 for avapritinib sensitivity, 0.81 for imatinib sensitivity. DL models predicted RFS, with hazard-ratios (HR) of 8.44 (95%CI 6.14–11.61) in the overall cohort and 4.74 (95%CI 3.34–6.74) in patients receiving adjuvant therapy. Prognostic performance was comparable to pathology-based scores, with highest discrimination in the overall cohort and in patients without adjuvant therapy (9.44, 95%CI (5.87–15.20)).

**Conclusion:** DL applied to WSIs enables prediction of molecular alterations, treatment sensitivity, and RFS in GIST, performing comparably to established risk scores across international cohorts, providing a baseline for future multimodal predictors.

**Highlights:**

- **Deep learning on histology predicts KIT and PDGFRA mutations in a large international cohort of GISTs from multiple centers**
- **Whole-slide image models stratify recurrence-free survival comparable to pathology-based risk scores**
- **Prognostic value of deep learning is preserved in adjuvant therapy subgroups, supporting treatment duration decisions**

Graphical abstract.
Overview of study design and dataset characteristics. (A) Multinational collection of WSIs from seven countries (Spain, France, Italy, Germany, the Netherlands, Poland, and Japan), followed by standard image preprocessing with the STAMP pipeline and clinical data preprocessing/standardization via the Grammar Data Curation framework. The workflow was divided into two main branches: (i) molecular mutation and treatment sensitivity prediction, and (ii) RFS prediction. Model performance was evaluated using AUROC and F1 score for classification tasks, and Kaplan–Meier survival curves with hazard ratios for RFS. Model explainability was assessed through heatmaps of WSIs and identification of top predictive tiles. (B) Summary of clinical dataset composition: proportion of cases receiving adjuvant therapy, tumor location distribution, mutation distribution at the exon level, and mutation distribution at the codon level.

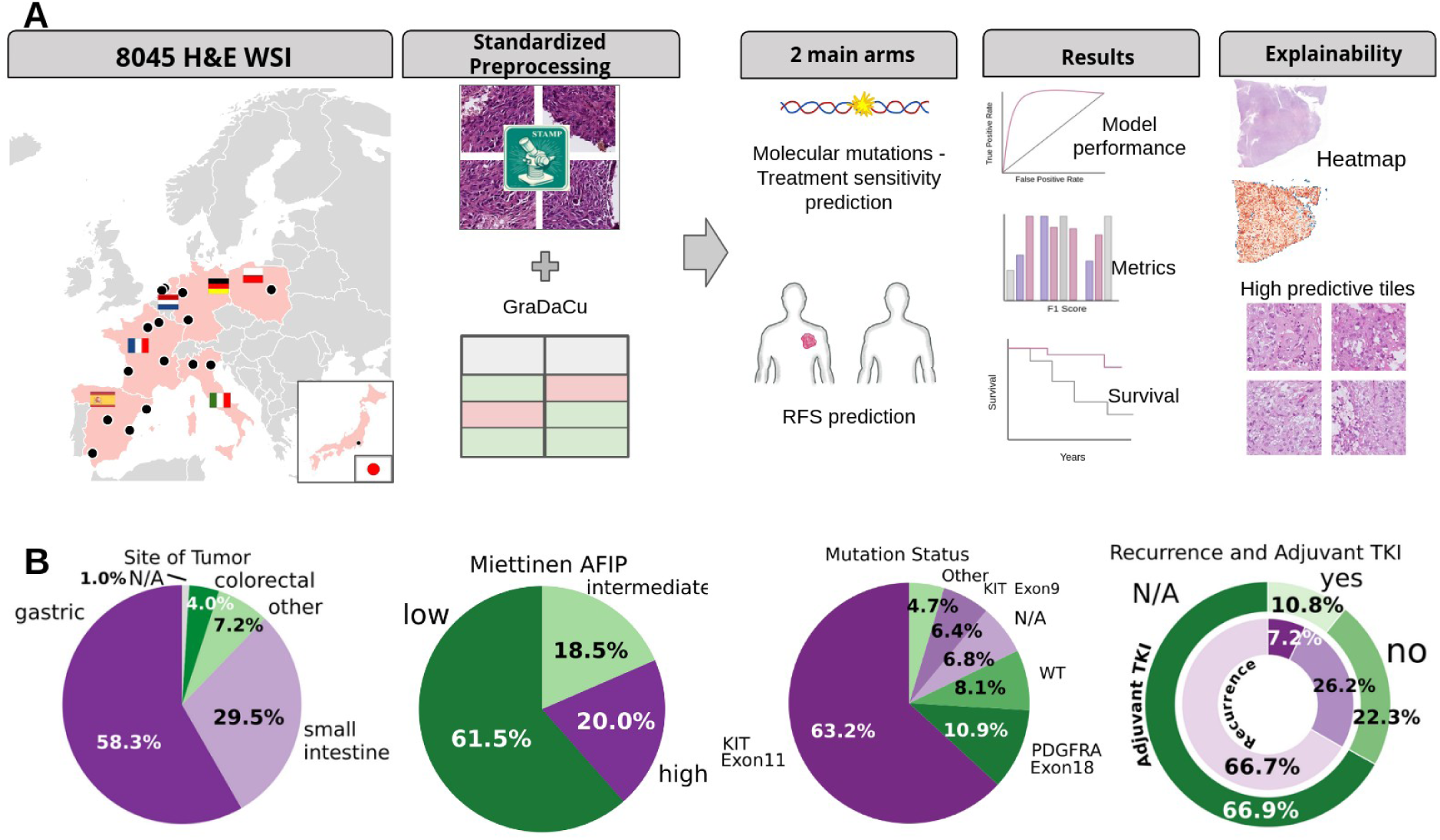

## Introduction

Gastrointestinal stromal tumors (GISTs) represent the most common mesenchymal neoplasms of the gastrointestinal tract^1^ with an annual incidence of 10–15 per million^2,3^, and a median age at diagnosis of approximately 60-65 years. Most originate in the stomach and small intestine,^4^ less commonly, they arise in the duodenum, rectum, colon, or esophagus.^5^

Molecularly, most GISTs harbor activating mutations in *KIT* (60-70%) or *PDGFRA* 10-15%^6,7^ both of which carry important prognostic and therapeutic implications. *KIT* exon 11 mutations, particularly at codons 557/558, are linked to aggressive behavior but predict strong benefit from adjuvant imatinib.^8,9^ *KIT* exon 9 mutations, (6-9%)^8^ typically require higher imatinib dosing for optimal efficacy in advanced phase, though not in adjuvant setting.^10^ *PDGFRA* exon 18 D842V mutations confer resistance to most tyrosine kinase inhibitors (TKIs) but respond to avapritinib^11,12,13^ Conversely, non-D842V *PDGFRA* exon 18 and exon 12 mutations remain generally imatinib-sensitive.^14^ Approximately 10-15% of GISTs are wild type (WT) for both genes, including SDH-deficient and NF1-associated subgroups, which derive limited benefit from current TKIs.^8,12^ Nevertheless, other mutations exist within WT GISTs, albeit with very low prevalence, such as those occurring in *BRAF* (sensitive to *BRAF* inhibitors) and in Neurotrophic Tyrosine Receptor Kinase (*NTRK*) family genes (sensitive to *NTRK* inhibitors). ^8,14^

Risk stratification is currently based on clinicopathological features such as tumor size, mitotic index, and site, with widely used systems including the NIH consensus criteria, the Armed Forces Institute of Pathology AFIP (Miettinen–Lasota) classification, and the modified Miettinen–Joensuu criteria (e.g., AFIP/Miettinen-Joensuu criteria).^9,15^ These systems, though widely used, are limited by inter-observer variability and reliance on subjective criteria like mitotic counting,^16^ underscoring the need for more objective prognostic tools.^17^

Deep learning (DL) applied to hematoxylin–eosin (H&E) whole-slide images (WSIs) for the extraction of prognostic and predictive morphologic features has emerged as a promising approach. Previous studies^11,12^ showed that DL can classify common GIST mutations with good accuracy, but limited cohort size and incomplete annotations raise concerns about generalizability.^18,19^ Such approaches should be considered complementary to, rather than replacements for, molecular testing, serving as an efficient prescreening tool to prioritize or rule out patients before molecular analysis.

To address these limitations, we assembled over 8,000 WSIs from 21 institutions in seven countries from Europe and Asia, with harmonized clinical, pathological, molecular, and follow-up data. Leveraging state-of-the-art foundation models and transformer architectures, we aimed to (i) predict *KIT* and *PDGFRA* mutation subtypes directly from WSIs, (ii) develop an integrated prognostic model for recurrence-free survival (RFS) incorporating adjuvant imatinib, and (iii) identify subgroups most likely to benefit from targeted therapy.

## Materials and Methods

### Patient Data Acquisition

We assembled a multi-center international cohort comprising 8,045 WSIs from 21 institutions across seven countries (France, Spain, the Netherlands, Germany, Italy, Poland, Japan; Supplementary Table 1). Comprehensive clinical, pathological, and molecular data were collected for each participant; Supplementary Table 2. Eligibility required a gastrointestinal tumor morphology consistent with GIST and positive immunostaining for *KIT* and/or DOG1. All samples were obtained from tumors that were chemotherapy– and TKI-naïve at the time of tissue collection. Collected variables included sex, age at diagnosis, relevant medical history (NF1, familial GIST, Carney Triad), type of sampling (resection, open biopsy, microbiopsy), tumor location, size, mitotic count, AFIP risk group, surgery, rupture, adjuvant TKI therapy, recurrence, and follow-up status. All participating centers were specialized in soft tissue tumors, and risk stratification according to AFIP/Miettinen-Joensuu was harmonized by an expert pathologist (JMC). Molecular data were curated at gene, exon, and codon levels.

### Image Processing and Deep Learning Techniques

#### Whole Slide digitization

Slides were stained with hematoxylin–eosin (HE) in most centers, while France applied hematoxylin–eosin–saffron (HES) Supplementary Table 1. WSIs were digitized at 40× magnification using local scanners (Hamamatsu NanoZoomer, 3DHISTECH Pannoramic, Aperio AT2). Despite minor inter-site differences, prior work^20^ has demonstrated cross-compatibility between HE and HES slides, supporting pooled analysis.

#### Data Preprocessing - WSI

WSIs were preprocessed with the STAMP pipeline^21^ for tessellation and background rejection; patches were saved as JPEGs and subsequently used for feature extraction with CONCH^22^, to train the downstream Vision Transformer (ViT) classification models. For each mutation, dichotomous variables were generated (positive for mutated, negative otherwise). Missing molecular tests were left blank per pipeline specifications. CONCH-derived features and the clinical or molecular data served as input for ViT^23^ classification model trained utilizing the default parameters as specified in the STAMP pipeline documentation.^21^

#### Data preprocessing - Clinical data

Clinical and molecular metadata were collected via a standardized template. Substantial inter-site heterogeneity in language, date formats, and mutation reporting required harmonization. Molecular information provided as nucleotide or protein formulas was standardized into categorical variables (gene/exon and codon level) using canonical Ensembl transcripts (KIT: NM_000222.2; PDGFRA: NM_006206.4). Key subtypes, including KIT exon 9 and KIT exon 11 ≥2 codon deletions/del–ins, were automatically identified using the *hgvs.parser* library.^24^

To ensure dataset consistency, we developed the Grammar Data Curation Tool (GraDaCu), a Python-based interface enforcing structured schema definitions, mandatory fields, and value constraints Supplementary Table 2. Iterative automated and manual checks ensured conformity before exporting a curated CSV for downstream analysis.

#### Mutation/clinical classification model

The overall dataset was first defined by samples from France, Germany, the Netherlands, and Japan, and was then partitioned 80:20 into training and internal validation cohorts, utilizing stratified sampling to preserve key variables. Conversely, samples sourced from Spain, Italy, and Poland were reserved entirely to form the independent external validation cohort. For mutation prediction, only resection samples were used for training, and the models were subsequently validated on two independent external cohorts: one composed exclusively of resections and another composed exclusively of biopsies, to test generalizability across sample types. AUC was assessed by bootstrap resampling (1,000 iterations) to derive 95% CIs. Models ranged from a baseline XGBoost using clinical data (age, sex, site, mitotic index) to DL approaches based on STAMP and COBRA^25^, the latter trained under three fine-tuning strategies. For explainability, the most predictive tiles were extracted and heatmaps were generated to visualize the precise regions of model focus.

#### Recurrence Free Survival Model

For recurrence-free survival (RFS), the DIGIST cohort was partitioned using the same geographical scheme as the mutation model (C1: all patients; C2: without adjuvant therapy; C3: with adjuvant therapy) excluding patients with a follow-up shorter than 12 months. DL scores (C1-C3) were developed and systematically compared against benchmark models based on mitotic count and AFIP criteria. Multivariable Cox regression with baseline covariates (age, sex, mutation, and adjuvant TKI for C1) was used to estimate hazard ratios (HRs) and then applied on internal and external validation cohorts. Model performance was assessed with Harrell’s C-index and integrated Brier score (IBS) and compared across cohorts with per-mutation testing. Kaplan–Meier curves were generated using training-median cut-points. A full methodological description is provided and represented in the Supplementary Methods and Supplementary Figure 1.

## Results

### Deep learning can predict mutational status in GIST directly from pathology slides

We evaluated whether foundation model-based DL pipelines could predict mutational status directly from H&E WSIs. Using a large multicentric, international cohort of surgically resected GISTs, we trained classifiers for 12 mutation categories and validated them on external cohorts.

DL models accurately predicted *KIT* and *PDGFRA* mutations, with AUCs ranging from 0.87 (95% CI 0.83–0.90) for *KIT* to 0.96 (95% CI 0.93–0.98) for *PDGFRA* in external validation (Table 1, Figure 1A). Performance was particularly strong for *PDGFRA* exon 18, including the clinically relevant D842V variant (AUC 0.93 [95% CI 0.90–0.95] external), and for *KIT* exon 11 (AUC 0.82 [95% CI 0.78–0.86]). By contrast, *KIT* exon 9, WT, and other rare mutations showed only moderate discrimination. We also evaluated the most predictive image tiles identified by the model for these classifications Figure 1B.

**Figure 1.**
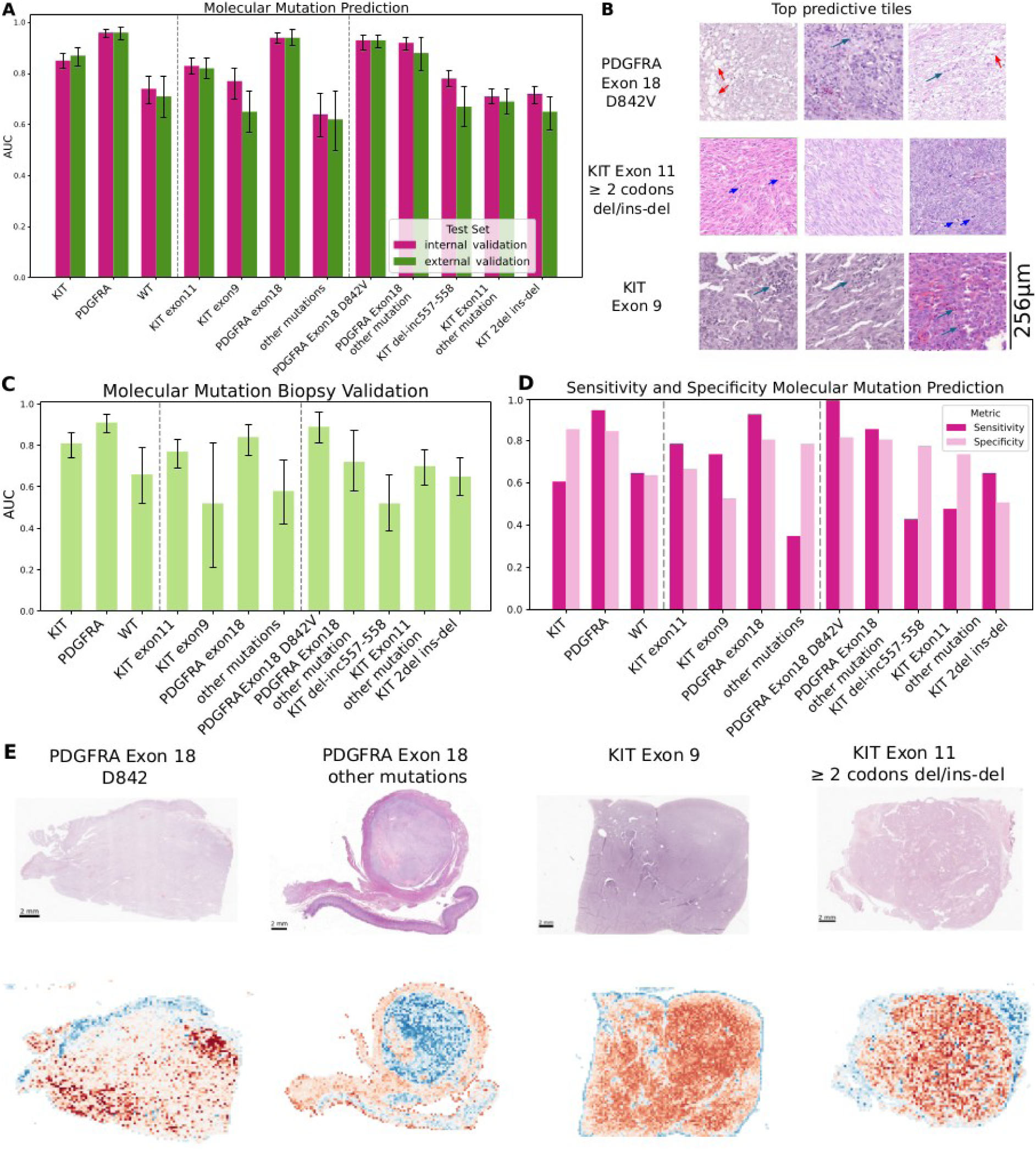
DL accurately predicts key molecular mutations in GIST from histopathology. (**A**) AUC with 95% confidence intervals for deep learning–based prediction of selected molecular mutations in the internal validation cohort (pink) and the external validation cohort (green). (**B**) Representative top-ranked predictive image tiles for three different mutations (PDGFRA Exon18 D842V, KIT exon 11 with 2 or mode codons deleted and KIT Exon9), as identified by the model’s attention score. Green arrows show lymphocytes for KIT exon9 and PDGFRA Exon18 D842V, red arrows show vacuolization of cells for PDGFRA Exon18 D842V and blue arrows show mitoses for KIT exon11 ≥ 2codons/del-ins (**C**) AUC with 95% confidence intervals for the same model shown in panel A when deployed on an external validation cohort consisting exclusively of biopsy specimens; results are reported for all selected mutations. (**D**) F1 scores for each mutation at the optimal classification threshold determined by Youden’s index (pink) and at a fixed sensitivity of 90% (light pink). (**E**) In the case of well-characterized mutations such as PDGFRA exon 18 D842V and KIT exon 9, the model focused on tumor regions, in line with reported morphological correlates. In other cases (PDGFRA Exon 18 other mutations), mucosal regions were also emphasized, likely due to vacuolated cells resembling PDGFRA-mutated morphology. Even for more specific variants, such as KIT exon 11 deletions or insertions, the model continued to highlight relevant tumor areas.

**Table 1.** Predictive performance of the DL models for selected molecular mutations in GIST. Results are reported as AUROC with 95% CI for the internal test set and the external validation set. For each mutation, the table also presents AUC (95% CI) values from external validation performed on biopsy-only samples. The WT group is defined by tumors with no mutations in both KIT and PDGFRA. The OTHER mutation group includes all mutations that are not categorized as KIT Exon 11, KIT Exon 9, PDGFRA Exon 18 or WT.

| **Molecular Mutation** | **Test set** | **AUC** | **CI lower** | **CI upper** |
| --- | --- | --- | --- | --- |
| KIT | internal test set | 0.85 | 0.82 | 0.88 |
| KIT | external validation | 0.87 | 0.83 | 0.9 |
| KIT | external validation biopsy only | 0.81 | 0.74 | 0.86 |
| PDGFRA | internal test set | 0.96 | 0.94 | 0.97 |
| PDGFRA | external validation | 0.96 | 0.93 | 0.98 |
| PDGFRA | external validation biopsy only | 0.91 | 0.86 | 0.95 |
| WT | internal test set | 0.74 | 0.68 | 0.79 |
| WT | external validation | 0.71 | 0.63 | 0.79 |
| WT | external validation biopsy only | 0.66 | 0.52 | 0.79 |
| KIT exon11 | internal test set | 0.83 | 0.8 | 0.86 |
| KIT exon11 | external validation | 0.82 | 0.78 | 0.86 |
| KIT exon11 | external validation biopsy only | 0.77 | 0.69 | 0.83 |
| KIT exon9 | internal test set | 0.77 | 0.7 | 0.82 |
| KIT exon9 | external validation | 0.65 | 0.57 | 0.73 |
| KIT exon9 | external validation biopsy only | 0.52 | 0.21 | 0.81 |
| PDGFRA exon18 | internal test set | 0.94 | 0.92 | 0.96 |
| PDGFRA exon18 | external validation | 0.94 | 0.91 | 0.97 |
| PDGFRA exon18 | external validation biopsy only | 0.84 | 0.75 | 0.9 |
| other mutations | internal test set | 0.64 | 0.55 | 0.72 |
| other mutations | external validation | 0.62 | 0.5 | 0.73 |
| other mutations | external validation biopsy only | 0.58 | 0.42 | 0.73 |
| PDGFRA Exon18 D842V | internal test set | 0.93 | 0.89 | 0.95 |
| PDGFRA Exon18 D842V | external validation | 0.93 | 0.9 | 0.95 |
| PDGFRA Exon18 D842V | external validation biopsy only | 0.89 | 0.81 | 0.96 |
| PDGFRA Exon18 other mutation | internal test set | 0.92 | 0.89 | 0.94 |
| PDGFRA Exon18 other mutation | external validation | 0.88 | 0.81 | 0.94 |
| PDGFRA Exon18 other mutation | external validation biopsy only | 0.72 | 0.58 | 0.87 |
| KIT del-inc557-558 | internal test set | 0.78 | 0.75 | 0.81 |
| KIT del-inc557-558 | external validation | 0.67 | 0.59 | 0.75 |
| KIT del-inc557-558 | external validation biopsy only | 0.52 | 0.39 | 0.66 |
| KIT Exon11 other mutation | internal test set | 0.71 | 0.68 | 0.74 |
| KIT Exon11 other mutation | external validation | 0.69 | 0.64 | 0.74 |
| KIT Exon11 other mutation | external validation biopsy only | 0.7 | 0.61 | 0.78 |
| KIT 2del ins-del | internal test set | 0.72 | 0.68 | 0.75 |
| KIT 2del ins-del | external validation | 0.65 | 0.58 | 0.71 |
| KIT 2del ins-del | external validation biopsy only | 0.65 | 0.56 | 0.74 |

In a separate biopsy cohort (n=223), performance declined, consistent with the smaller tissue area. *KIT* and *PDGFRA* remained well predicted with AUCs 0.81 (95% CI 0.74–0.86) and 0.91 (95% CI 0.86–0.95), respectively, while WT and rare variants were modest (AUCs ≤0.70; Figure 1C, Table 1)

External validation confirmed the robustness of the models (Figure 1D). *KIT* predictions reached sensitivity 0.61 and specificity 0.86, while *PDGFRA* achieved sensitivity 0.95 and specificity 0.85. The *PDGFRA* D842V variant reached perfect sensitivity (1.00) with specificity 0.82. Overall *KIT* exon 11 and *PDGFRA* predictions achieved the highest F1 scores (0.75–0.83). Full subgroup results are provided in Table 2.

**Table 2.** Sensitivity, specificity, and F1 score for molecular mutation prediction. Performance metrics of the DL model are reported for all molecular mutation categories in the external validation cohort. Values are presented for the optimal classification threshold determined by Youden’s index and for a fixed sensitivity of 90%.

| Molecular mutation | Sensitivity | Specificity | F1_opt | F1_sens90 |
| --- | --- | --- | --- | --- |
| KIT | 0.61 | 0.86 | 0.83 | 0.84 |
| PDGFRA | 0.95 | 0.85 | 0.78 | 0.29 |
| WT | 0.65 | 0.64 | 0.31 | 0.19 |
| KIT exon11 | 0.79 | 0.67 | 0.82 | 0.77 |
| KIT exon9 | 0.74 | 0.53 | 0.19 | 0.13 |
| PDGFRA exon18 | 0.93 | 0.81 | 0.75 | 0.26 |
| other mutations | 0.35 | 0.79 | 0.15 | 0.10 |
| KIT del-inc557-558 | 0.43 | 0.78 | 0.33 | 0.23 |
| KIT exon11 other mutation | 0.48 | 0.74 | 0.66 | 0.59 |
| PDGFRA exon18 D842V | 1.00 | 0.82 | 0.49 | 0.14 |
| PDGFRA exon18 other | 0.86 | 0.81 | 0.75 | 0.26 |
| KIT 2del ins-del | 0.65 | 0.51 | 0.42 | 0.36 |

Using a high-sensitivity threshold is appropriate for clinical pre-screening before confirmatory gold-standard testing. At 90% sensitivity, the number needed to test (NNTest) was 1.15, for *KIT*, 1.45 for *PDGFRA* and 7.0 for WT, corresponding to a relative reduction of 5%, 88%, and 30%, respectively.

We investigated whether a simple machine learning (ML) classifier based on five clinicopathological variables (sex, age, site of tumor, size of tumor, mitotic index) could predict mutations. Performance was lower than DL model. (Supplementary Table 3). These findings highlight that DL models trained on histopathology images outperform models based only on clinicopathological variables, underscoring their ability to capture morphological signals relevant to mutation status. In line with recent advances in DL-based molecular prediction, we also evaluated the COBRA workflow and observed results comparable to our baseline foundation model (Supplementary Table 4).

We conducted an explainability analysis to identify histopathological features associated with specific mutations and clinical outcomes. Heatmaps generated using Gradient-weighted Class Activation Mapping (Grad-CAM)^26^ (Figure 1E) highlighted the regions most relevant for prediction. When examining these regions and the most predictive tiles for different mutations, we observed that the *PDGFRA* exon 18 D842V (Figure 1E left) mutation was associated with epithelioid cell morphology, cytoplasmic vacuolization, myxoid stroma, and lymphoid infiltrates. In contrast, *KIT* exon 11 (Figure 1E right) mutations involving deletions of two or more codons were linked to mitotic activity and hyperchromasia, while *KIT* exon 9 mutations were associated with lymphoid infiltrates.

### Deep learning predicts clinically actionable alterations from pathology slides

We grouped mutations into three actionable categories: avapritinib-sensitive, imatinib-sensitive, and imatinib-dose-adjust as defined in Kong X et al. ^12^ (Supplementary Table 5) and trained DL models to predict each category from H&E slides (Figure 2A-B).

**Figure 2.**
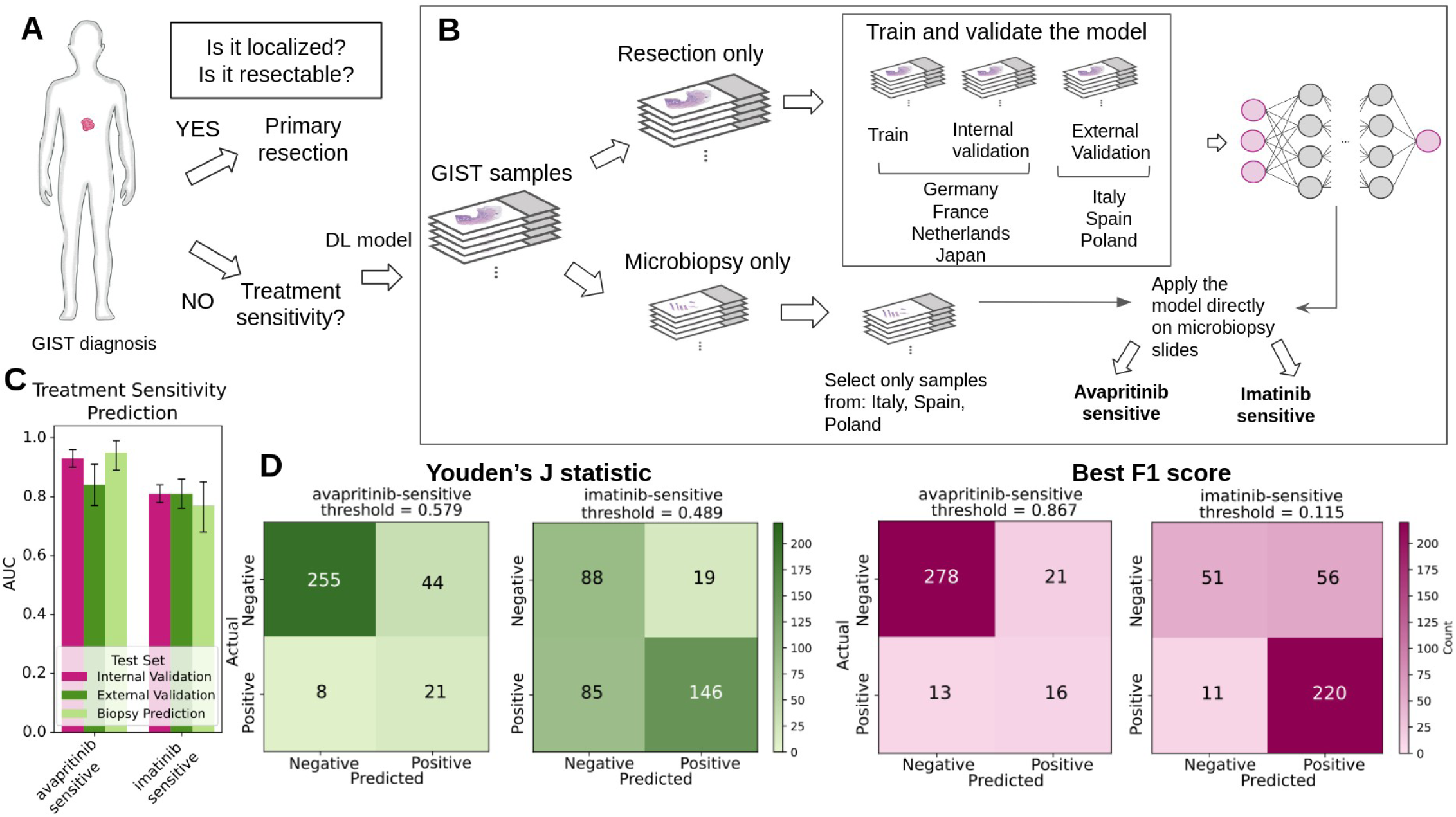
DL predicts treatment sensitivity and supports clinical decision-making in GIST. (**A**) Standard clinical workflow from initial diagnosis to treatment selection in patients with GIST. (**B**) Proposed integration of the deep learning model into the diagnostic–therapeutic pathway, enabling early triage of patients for further molecular analysis or tailored treatment selection. (**C**) AUC with 95% confidence intervals for deep learning–based prediction of treatment sensitivity, reported for the internal validation cohort (pink), external validation cohort (green), and an external biopsy-only validation cohort (light green). (**D**) Confusion matrices for avapritinib and imatinib sensitivity, shown at the optimal threshold by Youden’s index (green) and at the threshold yielding the highest F1 score (pink).

DL achieved strong predictive performance for avapritinib sensitivity (AUC 0.84 [95% CI: 0.77–0.91]) and consistent accuracy for imatinib sensitivity (AUC 0.81 [95% CI: 0.76–0.86]). By contrast, predictions for imatinib dose adjustment were weaker (AUCs ≤0.73; Table 3, Figure 2C).

**Table 3.** Predictive performance of the DL models for treatment sensitivity categories in GIST. Results are reported as AUROC with 95% CI for the internal validation set and the external validation set. For each treatment sensitivity category, the table also presents AUC (95% CI) values from external validation performed on biopsy-only samples.

| Treatment | Test set | AUC | CI_lower | CI_upper |
| --- | --- | --- | --- | --- |
| avapritinib-sensitive | internal test set | 0.93 | 0.9 | 0.96 |
| avapritinib-sensitive | external validation | 0.84 | 0.77 | 0.91 |
| avapritinib-sensitive | external validation biopsy only | 0.95 | 0.89 | 0.99 |
| imatinib-sensitive | internal test set | 0.81 | 0.78 | 0.84 |
| imatinib-sensitive | external validation | 0.81 | 0.76 | 0.86 |
| imatinib-sensitive | external validation biopsy only | 0.77 | 0.68 | 0.85 |
| imatinib-dose-adjust | internal test set | 0.78 | 0.71 | 0.84 |
| imatinib-dose-adjust | external validation | 0.73 | 0.64 | 0.82 |
| imatinib-dose-adjust | external validation biopsy only | 0.34 | 0.04 | 0.96 |

External validation confirmed predictive capacity, with avapritinib and imatinib sensitivities showing the most robust sensitivity/specificity balance, while dose-adjustment remained weaker (Table 4, Figure 2D). The “rule-out fraction” represents the proportion of patients who can be excluded from further testing at a given high-sensitivity threshold. At 90% sensitivity, the rule-out fractions were 0.50 for avapritinib-sensitive, 0.56 for imatinib-dose-adjust, and 0.24 for imatinib-sensitive for resections (Table 4).

**Table 4.**
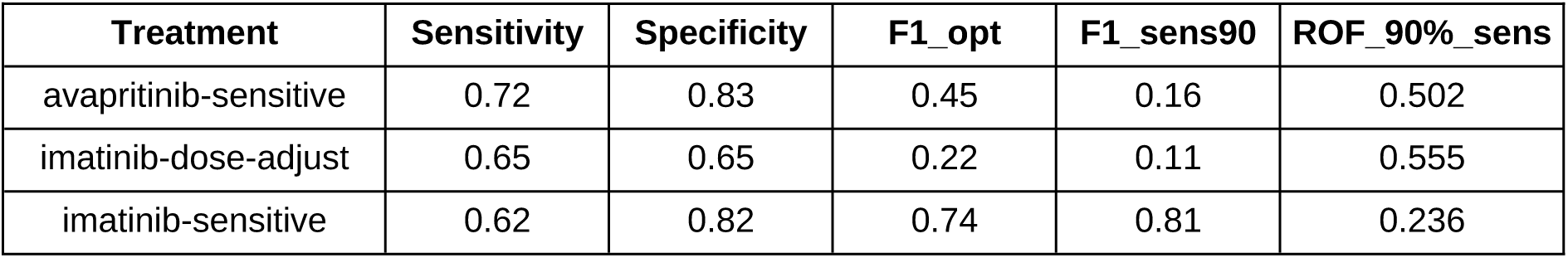
Sensitivity and specificity for treatment sensitivity prediction. DL model performance in the external validation cohort for avapritinib-sensitive, imatinib-dose-adjust, and imatinib-sensitive categories.

In comparison, a simple ML model trained only on clinicopathological variables performed poorly across all treatment categories (Supplementary Table 6), underscoring the added predictive value of DL.

### Deep learning predicts clinical outcomes in resected GIST

We next tested whether DL could directly predict RFS from H&E WSIs in the cohort C1 (Supplementary Table 7). The DL score strongly correlated with the mitotic count (p < 0.0001; Figure 3A) and provided significant prognostic information. When dichotomized at the training-set median, the DL score stratified patients into high– and low-risk groups with highly significant differences in RFS (HR of 8.44 (95% CI 6.14–11.61, p < 0.0001; Figure 3B Suppl. Table 8). In external validation, the simple DL model achieved a C-index of 0.67 (95% CI 0.59–0.75), comparable to the mitotic index (0.66, 95% CI 0.60–0.73) but lower than AFIP (0.80, 95% CI 0.74–0.88; Figure 3C; Table 5, Full results in Supplementary Table 9).

**Figure 3.**
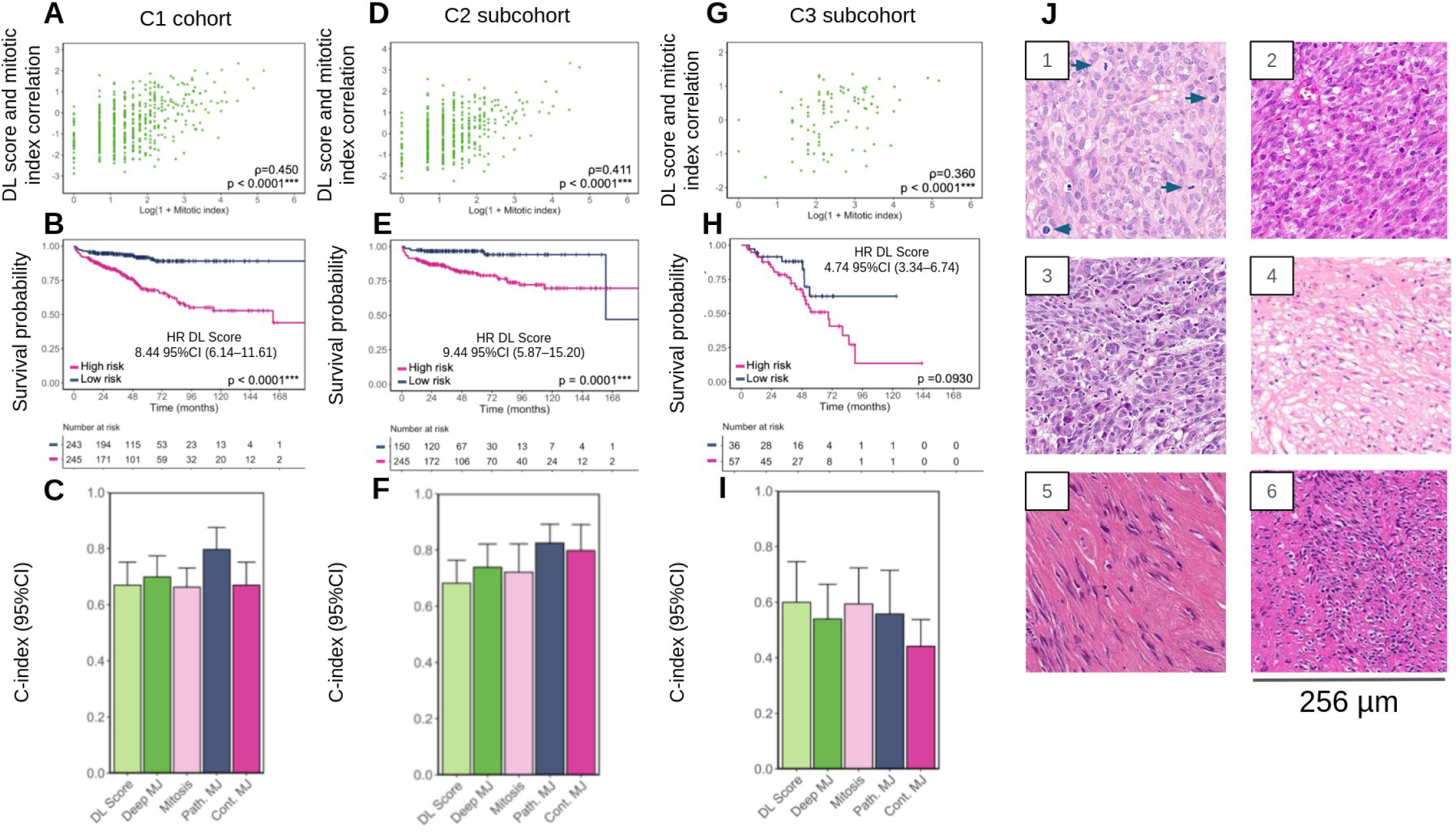
DL stratifies recurrence risk and complements clinical prognostic models in GIST. (**A–C**) Analyses in the overall C1 cohort: (**A**) correlations between the deep learning (DL) score and the mitotic index assessed by the Spearman rank test; (**B**) Kaplan–Meier curves for RFS according to the DL score (binarized at the median value of the C1 training dataset). (**C**) concordance indices (C-indices) with 95% confidence intervals (CIs) for five prognostic models: mitotic model (“mitosis”), simple DL model (“DL score”), pathological Miettinen–Joensuu model (“Path. MJ”), continuous Miettinen–Joensuu model (“Cont. MJ”), and deep Miettinen–Joensuu model (“Deep MJ”). (**D–F**) Corresponding analyses in the C2 subcohort (patients from C1 who did not receive adjuvant TKI therapy). (**G–I**) Corresponding analyses in the C3 subcohort (patients from C1 who received adjuvant TKI therapy). (**J**) Top representative tiles. Tiles 1, 2 and 3 from high risk tumors showed high cellularity with epithelioid cells, nuclear atypia and hyperchromasia. Tile 1 showed numerous mitoses (arrows). Tiles 4, 5 and 6 from low risk tumors showed low cellularity with spindle cells and fibrosis, no atypia and no hyperchromasia. *: p < 0.05; **: p < 0.005; ***: p < 0.001.

**Table 5.** Prognostic performance of mitotic index, deep learning (DL) score, and Miettinen–Joensuu (M.J.) risk models in cohorts C1–C3. C-index values (95% CI) are reported for each model. For the simple DL model, hazard ratios (HRs) for high versus low DL score (binarized at the median in the C1 training set) with 95% CI and corresponding p-values from univariable Cox regression are shown. Cohort C1 includes all patients, C2 comprises patients without adjuvant TKI therapy, and C3 those with adjuvant TKI therapy.

| Cohort | Model | C-index (95% CI) | HR high vs low DL (95% CI) | p-value |
| --- | --- | --- | --- | --- |
| **C1** | Mitotic model | 0.66 (0.60–0.73) | - | - |
|  | Simple DL model | 0.67 (0.59–0.75) | 8.44 (6.14–11.61) | <0.0001 |
|  | Pathological M.J. model | **0.80 (0.74–0.88)** | - | - |
|  | Deep M.J. model | 0.70 (0.63–0.77) | - | - |
| **C2** | Mitotic model | 0.72 (0.64–0.82) | - | - |
|  | Simple DL model | 0.68 (0.59–0.76) | 9.44 (5.87–15.20) | <0.0001 |
|  | Pathological M.J. model | **0.83 (0.76–0.89)** | - | - |
|  | Deep M.J. model | 0.74 (0.64–0.82) | - | - |
| **C3** | Mitotic model | 0.59 (0.46–0.72) | - | - |
|  | Simple DL model | **0.60 (0.44–0.75)** | 4.74 (3.34–6.74) | <0.0001 |
|  | Pathological M.J. model | 0.56 (0.41–0.71) | - | - |
|  | Deep M.J. model | 0.54 (0.38–0.66) | - | - |

In multivariable Cox analyses, the continuous DL score remained a strong independent predictor of RFS (HR 2.83, 95% CI 2.47–3.25, p < 0.0001; Supplementary Table 10), whereas the mitotic index did not (p = 0.175). This demonstrates that the DL score provides independent prognostic information beyond established histopathological criteria. Integrating DL features with Miettinen-Joensuu clinical variables modestly improved external discrimination (C-index 0.70, 95% CI 0.63–0.77; Table 5, Full results in Supplementary Table 9), but AFIP criteria continued to yield the best external performance.

Among 1,715 patients with mutation data, survival curves varied by subtype. *KIT* exon 11 deletions involving ≥2-codons deletions/del–ins and *KIT* exon 9 mutations had the poorest RFS (median ∼6–7 years), WT and *PDGFRA* exon 18 mutations had the most favorable outcomes (∼20 years), whereas other KIT exon 11 variants were intermediate (Supplementary Figure 2). For clarity, mutations were grouped as unfavorable (*KIT* exon 11 ≥2-codon deletions/del–ins, *KIT* exon 9) or favorable (WT, *PDGFRA* exon 18). Unfavorable mutations had median survival of ∼10 years versus ∼20 years for favorable, with HR 2.74 (95% CI 2.19–3.42, p < 0.0001;(Supplementary Table 8).

Overall, the DL score correlated with mitotic activity, stratified risk across molecular sub-types, and remained an independent predictor of prognosis, though its external performance was less robust than AFIP, likely reflecting cohort-level heterogeneity.

### Deep learning retains prognostic value across treatment subgroups

To test whether adjuvant therapy influenced prognostic performance, we stratified patients into those without adjuvant TKI (C2; Supplementary Table 11) and those with adjuvant TKI (C3; Supplementary Table 12). In both cohorts, the DL score correlated strongly with mitotic count (p < 0.0001; Figure 3D), supporting its biological relevance.

In C2, results mirrored those of the overall cohort C1. The DL model achieved a C-index of 0.68 (95% CI 0.59–0.76) in external validation, modestly lower than the mitotic index (0.72, 95% CI 0.64–0.82) and Deep Miettinen–Joensuu (0.74, 95% CI 0.64–0.82) (Figure 3F, Table 5, Full results in Supplementary Table 9). Nevertheless, Kaplan–Meier curves showed clear separation between DL high– and low-risk groups (log-rank p < 0.0001; Figure 3E), with Cox regression confirming a strong prognostic effect (HR 9.44, 95% CI 5.87–15.20, p < 0.0001; Supplementary Table 8).

In C3, the DL model achieved a C-index of 0.60 (95% CI 0.44–0.75), comparable to the mitotic index (0.59, 95% CI 0.46–0.72) and superior to AFIP (0.56, 95% CI 0.41–0.71) (Figure 3I, Table 5, Supplementary Table 9). In Kaplan–Meier analysis, the survival difference between DL high– and low-risk groups did not reach statistical significance (log-rank p = 0.093; Figure 3H). However, Cox regression still demonstrated a strong prognostic effect (HR 4.74, 95% CI 3.34–6.74, p < 0.0001; Supplementary Table 8). Adding DL to AFIP did not further improve performance, despite the multivariable cox regression demonstrating a strong, independent prognostic effect.

When focusing on patients with imatinib-sensitive mutations, the DL score provided particularly sharp stratification for both RFS and OS. Among patients with a high AFIP score but not receiving adjuvant TKI, DL low-risk cases had significantly better RFS and OS (log-rank p = 0.0007 and p = 0.0333; Supplementary Figure 3A-B, Supplementary Table 13). Conversely, among patients with high AFIP score receiving adjuvant TKI, the DL score separated individuals into clinically meaningful subgroups for both RFS and OS (log-rank p < 0.0001 for both; Supplementary Figure 3BC-D, Supplementary Table 13).

Comparisons across treatment arms further underscored this effect. DL low-risk patients without adjuvant TKI had higher recurrence and worse OS compared with those receiving therapy, suggesting that a time-limited treatment (∼36 months) may be sufficient. By contrast, DL high-risk patients had poor outcomes regardless of treatment, with median recurrence time (∼60 months) exceeding the median treatment duration (∼35 months), supporting the potential need for prolonged or lifelong TKI therapy (Supplementary Figure 3, Supplementary Table 13). Representative predictive tiles are shown in Figure 3J.

Overall, the DL score retained prognostic value across treatment subgroups. While discrimination was modestly lower than some traditional criteria and KM separation was weaker in C3, Cox regression consistently confirmed significance. Importantly, subgroup analyses suggest that the DL score could help guide treatment duration by identifying low-risk patients for time-limited therapy and high-risk patients who may require prolonged or lifelong treatment, consistently with a recently published study showing longer duration benefit in high risk patients^27^.

## Discussion

We conducted a comprehensive, multicenter international analysis to predict molecular status and stratify recurrence risk in GISTs directly from routinely available pathology slides. Our findings demonstrate that DL models, trained on a large and diverse cohort of over 8,000 samples from 21 institutions across 7 countries, can accurately predict the most common and clinically actionable *KIT* and *PDGFRA* mutation subtypes directly from native tumor morphology. Crucially, this study validates that a DL-derived score provides strong, independent prognostic information for RFS. This work addresses a critical limitation of previous studies^11^ like small cohort size and limited generalizability by confirming the robustness of DL across varied international cohorts and demonstrating its utility as a non-invasive, complementary pre-screening tool to augment traditional GIST risk stratification and potentially guide adjuvant therapy duration.

For molecular prediction, our models achieved high accuracy for *KIT* and *PDGFRA* mutations, including near-perfect performance for *PDGFRA* exon 18 D842V (genotype conferring primary resistance to imatinib)^2,8^ and strong discrimination of *KIT* exon 11 variants. By contrast, *KIT* exon 9, WT, and rare subtypes were predicted with only moderate accuracy, reflecting both biological heterogeneity and smaller sample sizes. Importantly, DL outperformed models trained solely on clinical variables, underscoring that morphological features capture rich genetic information. The robustness of DL predictions across both resections and biopsies supports their potential clinical utility, although performance on biopsy specimens was slightly lower, consistent with the reduced amount of tissue available for analysis. The clinical relevance of molecular prediction is particularly significant for populations facing economic constraints, such as those in Low– and Middle-Income Countries, where resource-intensive sequencing diagnostics are often unavailable or prohibitively expensive as highlighted in ^28^. The DL approach provides a cost-effective triage strategy to streamline case selection for confirmatory molecular testing.

When grouped into clinically actionable categories, DL predictions distinguished avapritinib-and imatinib-sensitive tumors with high accuracy, while dose-adjustment was less reliable. The ability to compute “rule-out fractions” at high sensitivity suggests DL could serve as a rapid pre-screening tool, helping prioritize confirmatory testing and triaging cases in re-source-limited settings.

For outcome prediction, the DL score correlated strongly with mitotic activity and stratified patients into distinct risk groups. In the overall cohort, DL achieved C-indices similar to the mitotic index but lower than AFIP, but remained independently prognostic in multivariable models. Notably, the mitotic index lost significance when modeled alongside DL, indicating that DL captures its prognostic signal more reproducibly. Mutation grouping into unfavorable versus favorable provided a clinically meaningful simplification, consistent with prior reports^29^, and illustrates how DL-derived scores can be integrated with molecular features.

Stratification by treatment status provided further insights: in patients without adjuvant TKI, DL discrimination was modestly lower than classical criteria but still provided strong risk separation. In patients receiving adjuvant therapy, DL performed comparably to mitotic index and better than AFIP. While Kaplan-Meier curves showed weaker separation in external validation, Cox regression confirmed the DL score’s prognostic significance. Adjuvant imatinib is known to improve relapse-free survival in high-risk GIST, yet only one trial has demonstrated an overall survival benefit, favoring 3 years of treatment over 1 year.^9^ Therefore, the specific patient subgroup deriving the greatest benefit from adjuvant imatinib therapy is still uncertain, as is the optimal duration of treatment. In our study, we studied 628 high-risk patients according to the Miettinen-Joensuu classification and with a mutation sensitive to imatinib. In this group of patients 445 received adjuvant imatinib and 183 did not. The DL Miettinen-Joensuu model split both subgroups of patients into low and high DL Miettinen-Joensuu with a significant better recurrence and overall-free survival in DL low risk as compared to the DL high risk in both groups without and with adjuvant imatinib. Moreover, results suggested that time-limited (36 months) treatment could be sufficient in DL low risk although results on DL high risk support a potential benefit from prolonged or lifelong therapy.

Finally, explainability analyses conducted with top predictive tiles and heatmaps revealed that DL models focused on tumor regions and morphological features consistent with established pathological correlates, as also observed by Fu et al.^11^ High cellular density and mitotic activity characterized high-risk tumors, while PDGFRA D842V mutations displayed epithelioid or mixed morphology with cytoplasmic vacuolization, myxoid stroma, and lymphoid infiltration; KIT 557/558 deletions showed mitotic activity. These findings demonstrate that DL accurately captures morphological hallmarks recognized by pathologists, enhancing interpretability and clinical relevance.

Several limitations should be acknowledged. Despite the large dataset, rare genotypes such as *KIT* exon 9, SDH-deficient, or NF1 GISTs remain underrepresented, limiting performance estimates. Future studies should enrich these cohorts and validate generalizability prospectively. Moreover, epidemiological and pathological variations in GIST across populations suggest that our dataset may not fully represent the spectrum of Asian disease patterns, as Asian cases were underrepresented in this study (475 samples) compared with the predominantly European cohort. Finally, our models relied on histopathology but integration with radiology, genomic, and clinical data through multimodal architectures may enhance performance.

In conclusion, DL applied to WSIs accurately predicts molecular alterations, treatment sensitivity, and recurrence risk in GIST. While AFIP remains a strong comparator, DL provides independent prognostic information, identifies clinically actionable alterations, and refines treatment stratification. By combining predictive accuracy with interpretability, DL has the potential to complement molecular testing, guide adjuvant therapy decisions and support more personalized management of GIST.

## Data Availability

All data produced in the present work are contained in the manuscript

## Acknowledgments

The authors gratefully acknowledge the GWK support for funding this project by providing computing time through the Center for Information Services and HPC (ZIH) at TU Dresden.

The authors gratefully acknowledge the Gauss Centre for Supercomputing e.V. (www.gauss-centre.eu) for funding this project by providing computing time through the John von Neumann Institute for Computing (NIC) on the GCS Supercomputer JUWELS at Jülich Supercomputing Centre (JSC)

## Ethics statement

This study was performed in accordance with the Declaration of Helsinki. This study is a retrospective analysis of scanned images of anonymized tissue samples of various cohorts of cancer patients. Data were collected and anonymized and ethical approval was obtained. The overall analysis was approved by the Ethics board of the Medical Faculty of Technical University Dresden under the Approval ID: BO-EK-444102022_2.

## Author contributions

JMC and JNK conceptualized the study. AB and ZIC curated the source data. AB and VLL developed the source codes for the analysis and conducted the experiments. FW developed the source code for the Grammar Data Curation tool. AB, VLL, ZIC and MG interpreted the data, AB, VLL, ZIC, JMC, JNK and AC wrote the first draft of the manuscript. SWL, LV, PS, FLL, ML, PR, JMS, NS, HB, IM, SB, SN, EM, CA, FG, GT, AHG, GCJ, MC, SLP, REP, IS, EK, LB, EW, AM, JYB, PC, AV, YY, AG, HH, AG, TN, OB, JFE, CN, PH, CC, JJ, JVMGB, HG, ASC, MJD, JB, NL, AL, JYB, AI, AC provided images and data. All authors revised the manuscript draft, contributed to the interpretation of the data and agreed to the submission of this paper.

## Declaration of Generative AI and AI-assisted technologies in the writing process

During the preparation of this work the author(s) used Google Gemini 2.5, ChatGPT 4 and ChatGPT 5 in order to improve grammar, sentence structure, and overall clarity. After using this tool/service, the author(s) reviewed and edited the content as needed and take(s) full responsibility for the content of the publication.

## Data and Code availability

The only original code from this study is the Grammar Curation Tool, available at https://github.com/KatherLab/GraDaCu under MIT license. The code was developed specifically for this study and does not include re-used components from previously published repositories or software. All other analyses used previously published code.

## Supplementary Methods

For the analysis of RFS, the DIGIST cohort was partitioned using the same geographical split and the same ratio for defining training, internal and external validation cohorts as for the molecular prediction model. (Supplementary Figure 1 A).

Different DL models were trained on C1, C2 and C3 training sets to address the prognostic tasks and were subsequently named DL scores C1, C2 and C3. For each cohort, a same pipeline was employed to evaluate the prognostic value of the scores and their added value compared to the mitotic count alone and combined with the other established prognostic variables from the AFIP criteria (i.e., tumor size, location and rupture) (Supplementary Figure SF1 B). Age, sex, and mutational status (as well as adjuvant TKI therapy for the C1 cohort) were systematically included as baseline covariates in all models to account for their poten-tial confounding effect and to ensure consistent risk adjustment across model comparisons. Overall, five models were systematically trained on training C1, training C2 and training C3 relying on the (i) DL score (named simple DL model), (ii) DL score combined with tumor size, location and rupture (named deep Miettinen-Joensuu model), (iii) Mitotic count (named mi-totic model), (iv) AFIP criteria provided by the pathologists (named pathological Miettinen-Joensuu model), and (v) mitotic count combined with tumor size, location and rupture (named continuous Miettinen-Joensuu model). The performance of these DL-based models were systematically compared against the three other benchmark models providing insights into whether AI-driven approaches can enhance or complement existing prognostic tools.

Univariable survival analysis including log-rank tests and Cox regressions was first per-formed in each training cohort on the variables entered in the models.

Afterwards, the multivariable models were trained using the Cox proportional hazard ratio (HR) model in the training cohorts and applied on the internal and external validation cohorts. Multivariable HRs with their 95% CI were calculated in the training cohorts. The performances of the models were estimated using Harrell concordance index (C-index, which assesses the discriminative ability of the models) and integrated Brier score (IBS, combined with prediction error curves, which quantifies overall prediction accuracy and calibration over time) over the first 10 years following resection with 95% CIs. In the training cohorts, the model performances were evaluated using 5-fold cross-validation, with evaluation performed on the out-of-fold samples (Supplementary Figure 1 B).

Model performances were then compared in the internal and external validation cohorts using permutation tests, where predicted risks were randomly swapped (with 1000 random label exchanges) between models to generate the null distribution of differences in c-index and IBS. P-values were computed as the proportion of permuted differences exceeding the observed one. Kaplan-Meier curves for RFS in each patient group were drawn after dichotomizing the DL scores according to their median in the training cohorts.

## Supplementary Figures

**Supplementary Figure 1.**
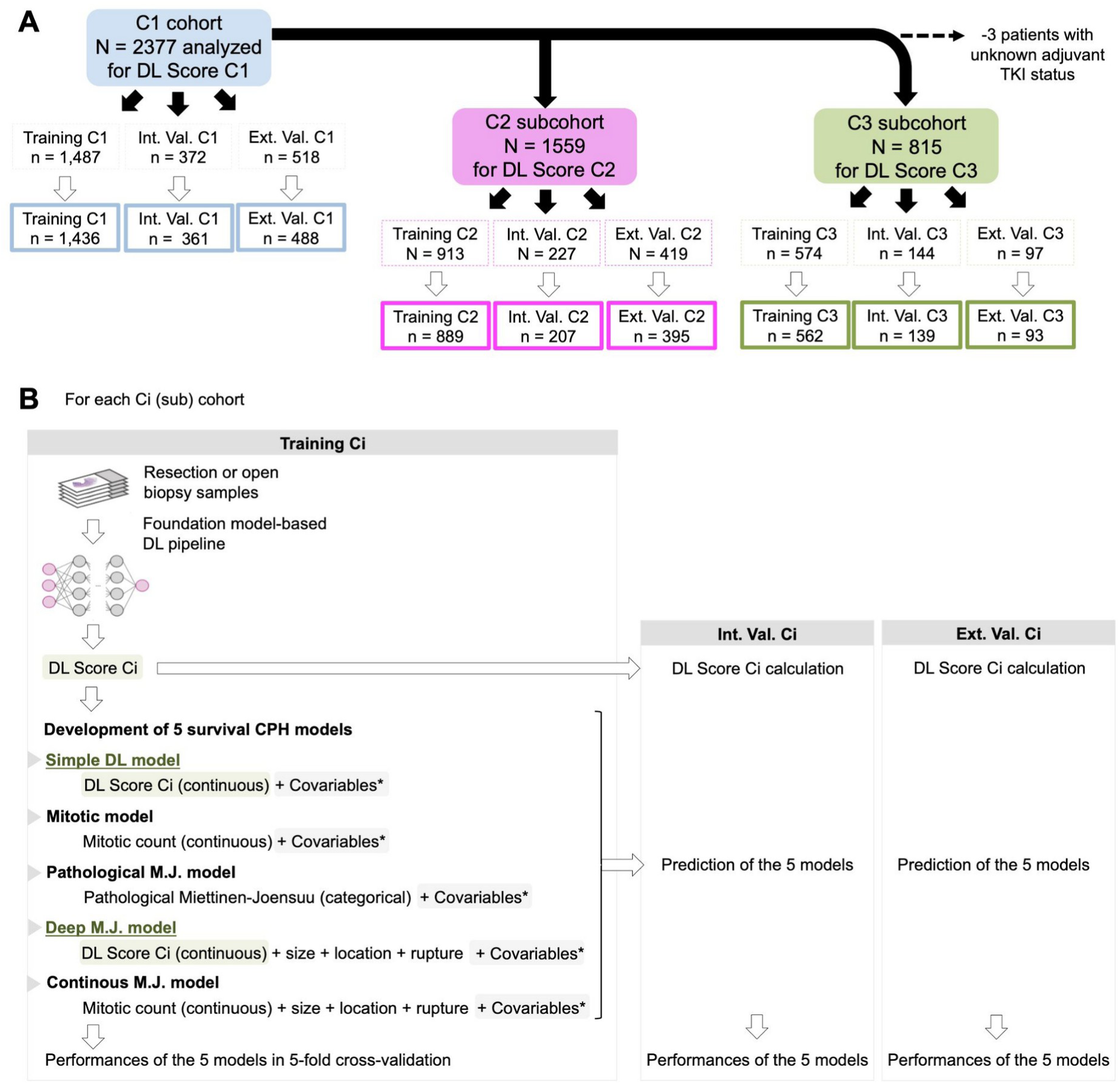
Statistical pipeline for recurrence-free survival (RFS) analysis. (A) Data partitioning. Boxes with light dashed contours corresponded to the patients with available DL Scores while boxes with solid thick contours to the patient included in the survival analysis. (**B**) Methodological approach performed in C1, C2 and C3 to obtain deep learning (DL) scores and to develop survival models in the Training cohorts using the Cox proportional hazard (CPH) algorithm and to compare them. Other abbreviations: ext.val: external validation, int. val: internal validation, M.J.: Miettinen-Joensuu. *Covariables were: age, sex, adjuvant TKI therapy and mutational status for C1; and age, sex and mutational status for C2 and C3.

**Supplementary Figure 2.**
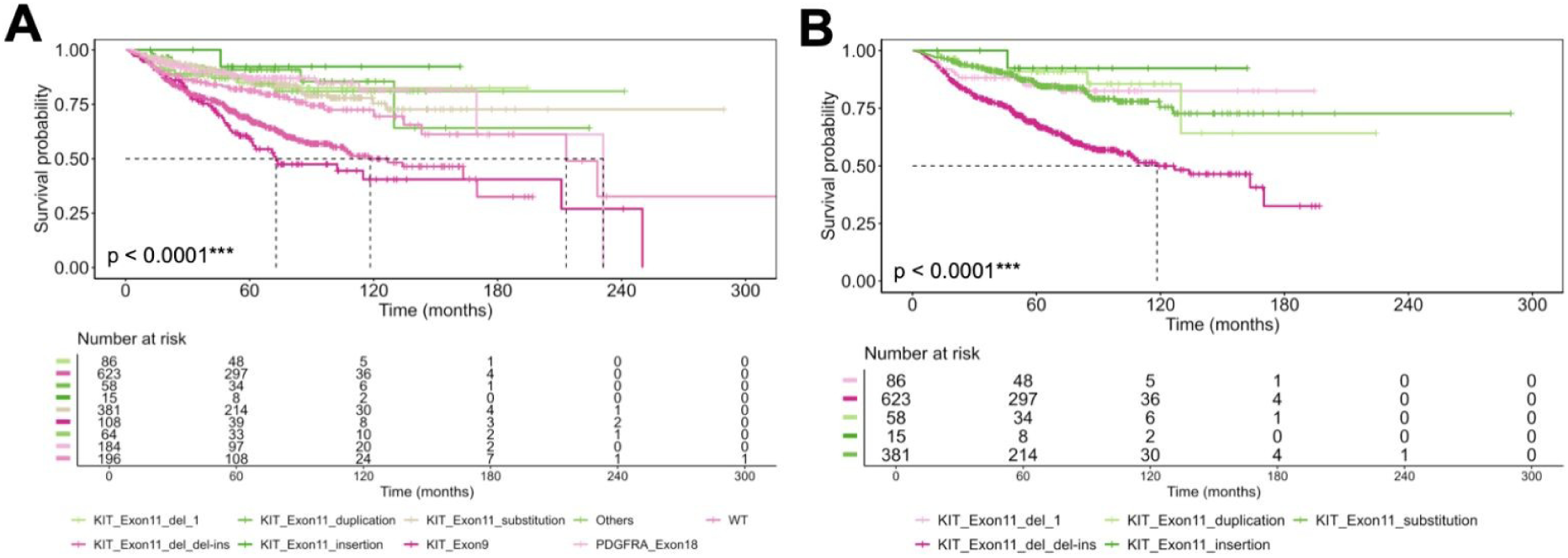
Associations between recurrence-free survival (RFS) and mutations. Kaplan-Meier curves in the whole C1 cohort (unfiltered) for all mutations (**A**) and for KIT mutations (**B**) *: p < 0.05; **: p < 0.005; ***: p < 0.001. Tests are log-rank tests.

**Supplementary Figure 3.**
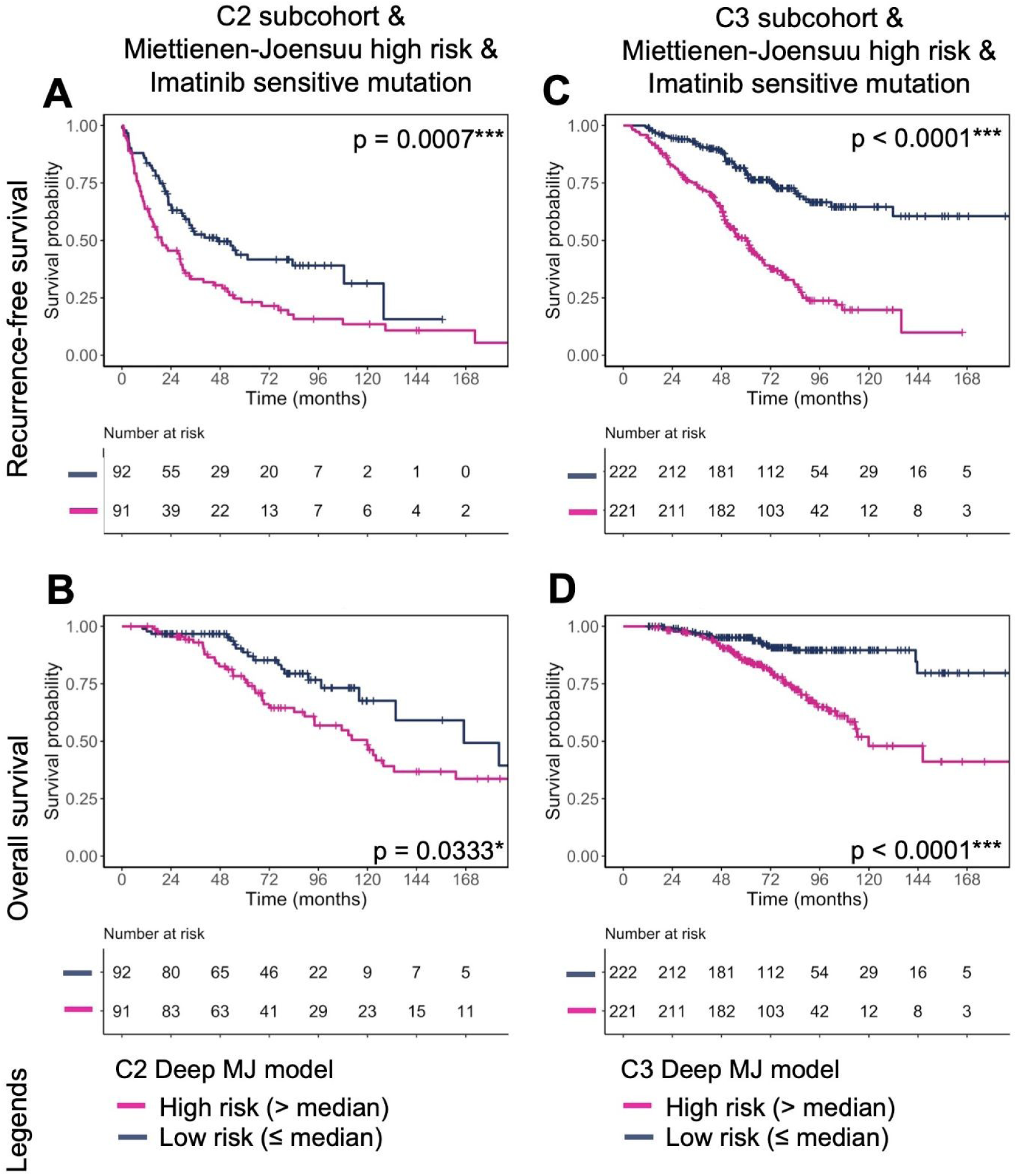
Kaplan-Meier curves for recurrence-free survival (RFS) and overall survival (OS) in the subgroup of C2 and C3 patients with high risk score according to pathological Miettienen-Joensuu scoring system and Imatinib sensitive mutations depending on the deep Miettinen-Joensuu models. Kaplan-Meier curves for RFS (**A**) and OS (**B**) depending on the deep Miettinen-Joensuu model employing the C2 DL Score in this subgroup of the C2 subcohort. Kaplan-Meier curves for RFS (**C**) and OS (**D**) depending on the deep Miettinen-Joensuu model employing the C3 DL Score in this subgroup of the C3 subcohort. *: p < 0.05; ***: p < 0.001. Tests are log-rank tests.

## Supplementary Tables

**Supplementary Table 1.**
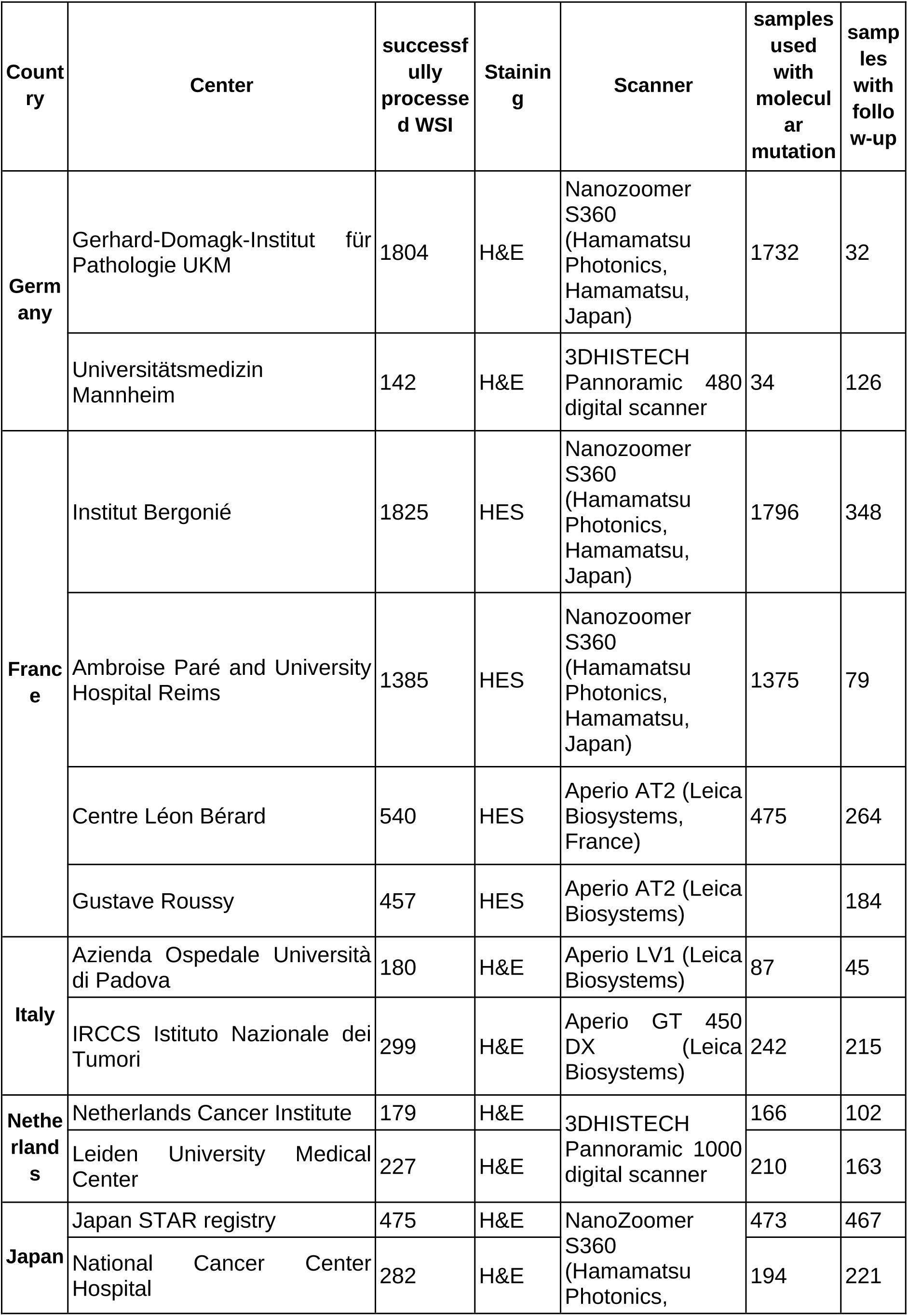
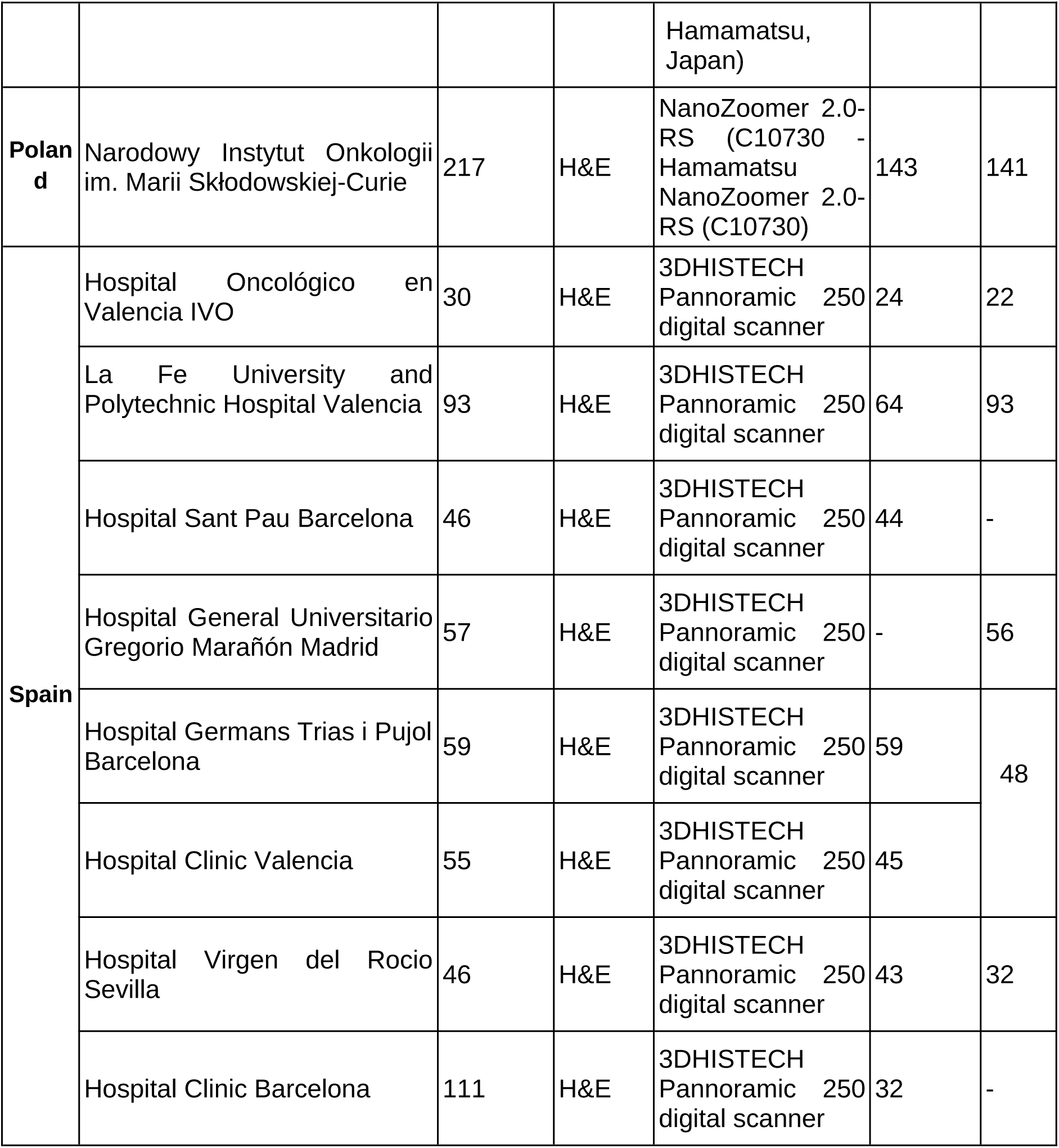
Sample collection and processing across participating centers. For each center, the table reports (1) the number of successfully processed slides (defined as slides from which features were correctly extracted), (2) the number of samples per cohort with available molecular mutation data, and (3) the number of samples with complete information required for inclusion in the RFS prediction analysis.

**Supplementary table 2.**
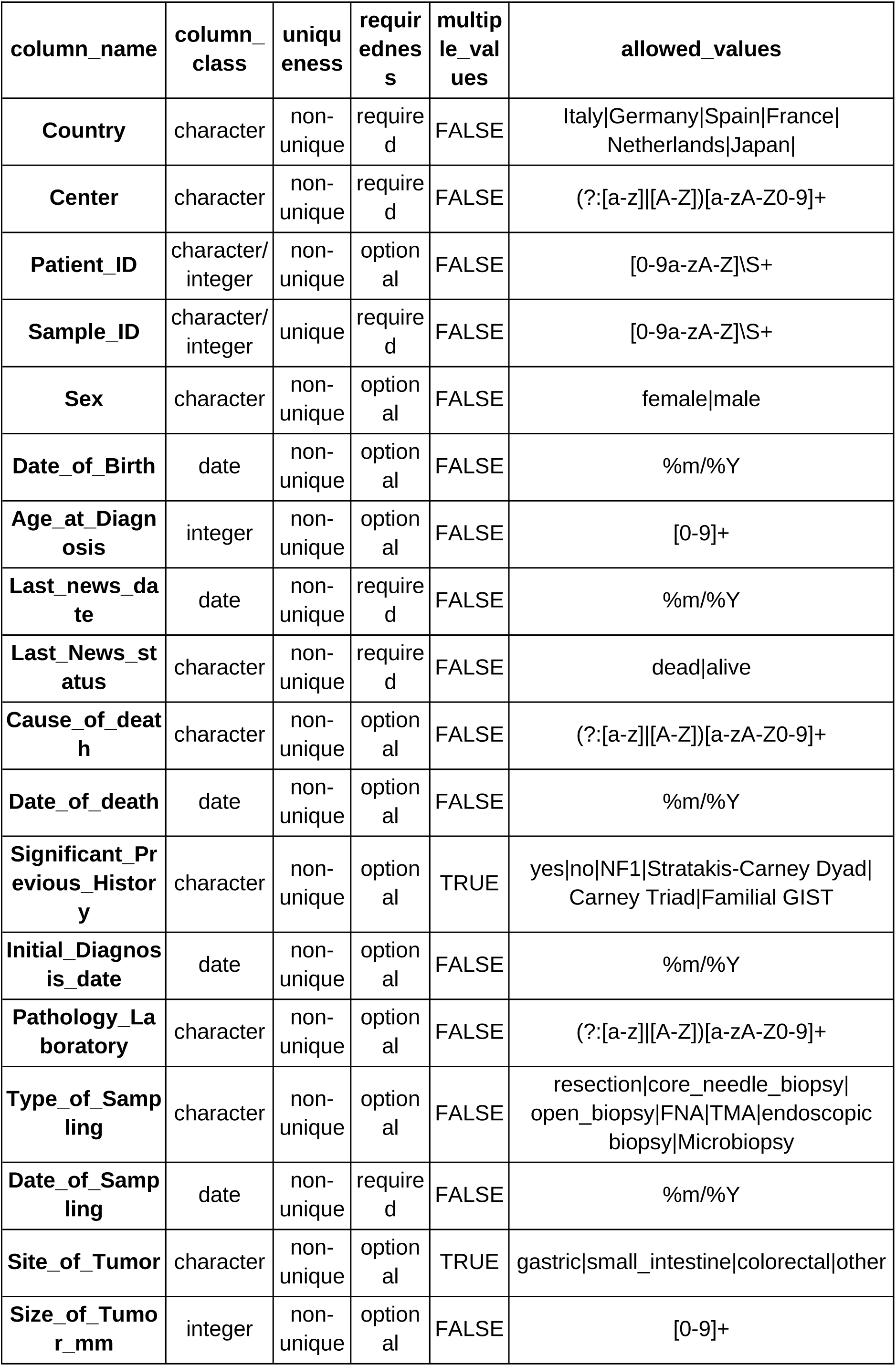
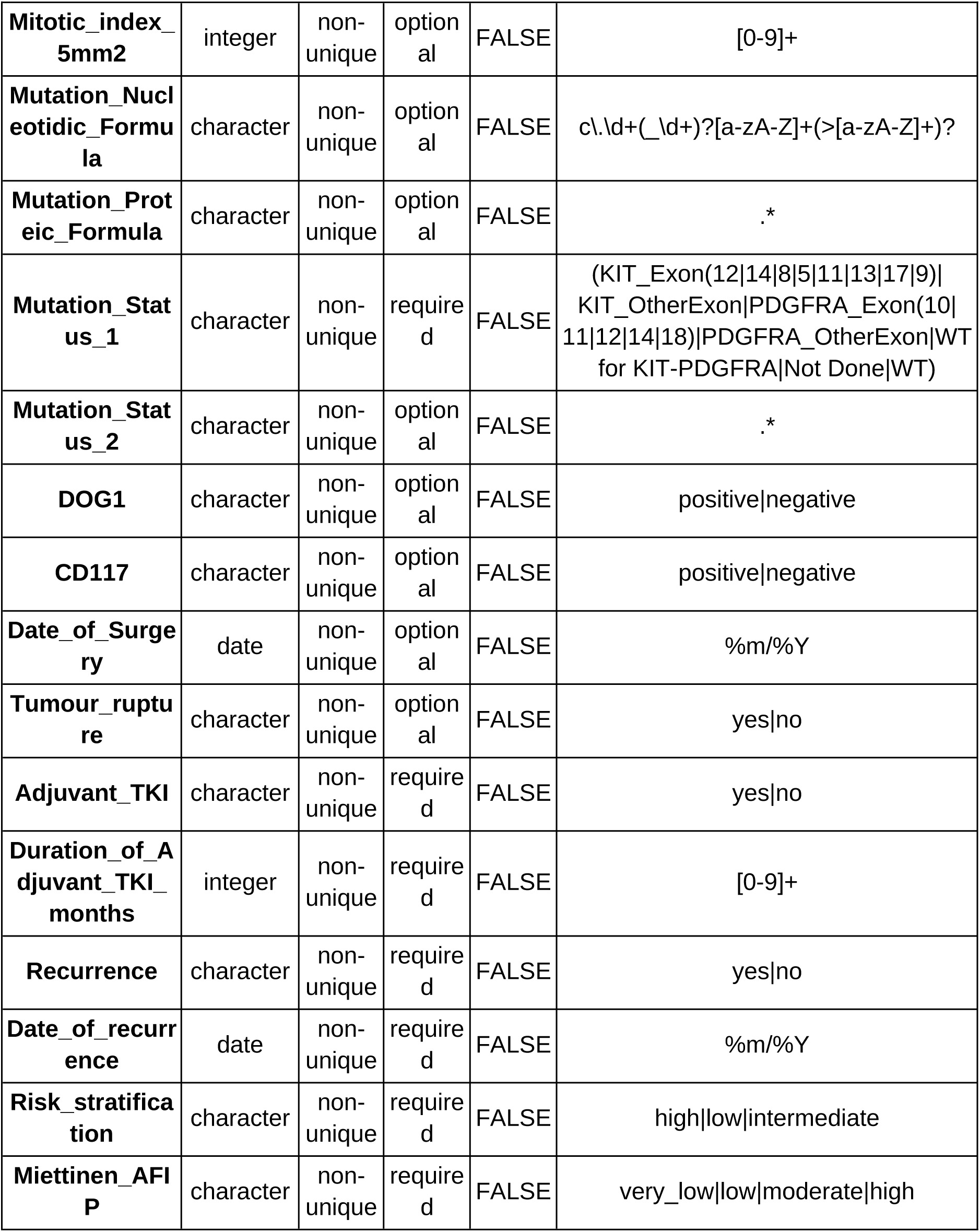
Template used for clinical data collection and harmonization across centers. The table defines all potential data fields included in the study, specifying the expected data type (*column_class*), uniqueness within the dataset, requiredness for analysis, allowance for multiple values, and permitted or regular-expression–defined entries. This schema served as both a reference for data collection and as the grammar underlying the automated Grammar Data Curation Tool.

**Supplementary Table 3.** Performance of ML models trained exclusively on clinical variables (sex, age, tumor location, mitotic index, and tumor size) for prediction of individual molecular mutations in GIST. For each mutation, the table reports accuracy, precision, recall, and the AUC with 95% CI.

| Molecular Mutation | Accuracy | Precision | Recall | F1 | AUC | CI_lower | CI_upper |
| --- | --- | --- | --- | --- | --- | --- | --- |
| WT | 0.78 | 0.21 | 0.41 | 0.27 | 0.64 | 0.58 | 0.68 |
| KIT exon11 | 0.61 | 0.61 | 1.00 | 0.76 | 0.53 | 0.48 | 0.58 |
| KIT exon9 | 0.62 | 0.17 | 0.90 | 0.28 | 0.80 | 0.76 | 0.84 |
| PDGFRA exon18 | 0.68 | 0.29 | 0.77 | 0.42 | 0.71 | 0.66 | 0.75 |
| other mutations | 0.05 | 0.05 | 1.00 | 0.10 | 0.62 | 0.56 | 0.68 |
| PDGFRA exon18 D842V | 0.08 | 0.08 | 1.00 | 0.15 | 0.78 | 0.73 | 0.83 |
| PDGFRA exon18 other | 0.92 | 0.20 | 0.19 | 0.20 | 0.68 | 0.63 | 0.74 |
| KIT del-inc557-558 | 0.70 | 0.18 | 0.37 | 0.24 | 0.55 | 0.48 | 0.61 |
| KIT exon11 other mutation | 0.45 | 0.43 | 0.95 | 0.59 | 0.53 | 0.47 | 0.59 |
| KIT | 0.73 | 0.73 | 1.00 | 0.84 | 0.56 | 0.51 | 0.61 |
| PDGFRA | 0.17 | 0.17 | 1.00 | 0.29 | 0.70 | 0.65 | 0.76 |
| KIT 2del ins-del | 0.73 | 0.38 | 0.42 | 0.40 | 0.64 | 0.59 | 0.70 |

**Supplementary Table 4.**
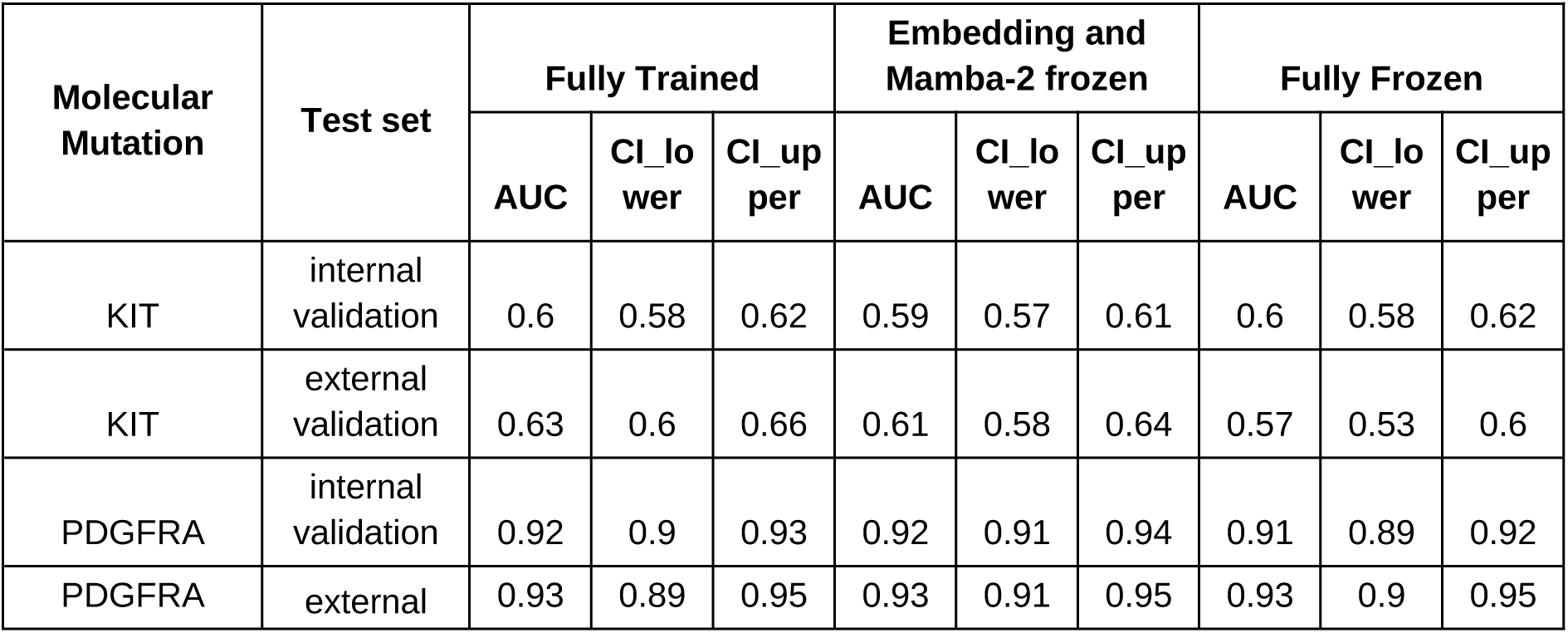
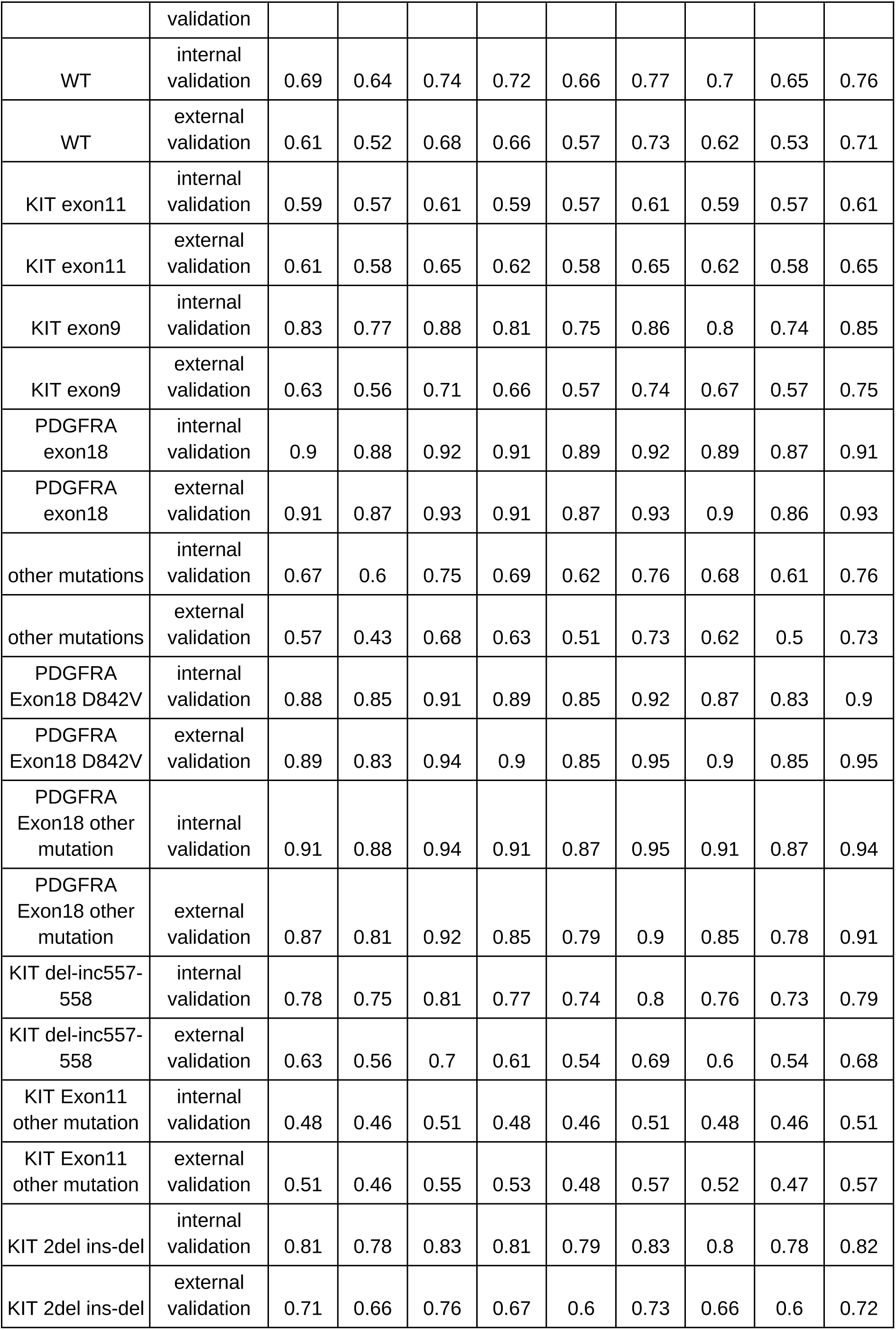
Predictive performance of the COBRA model for molecular mutations in GIST, reported as AUC with 95% CI for both the internal validation and the external validation. Results are presented for three training setups: fully trained, embedding and Mamba-2 layer frozen, and fully frozen.

**Supplementary Table 5.**
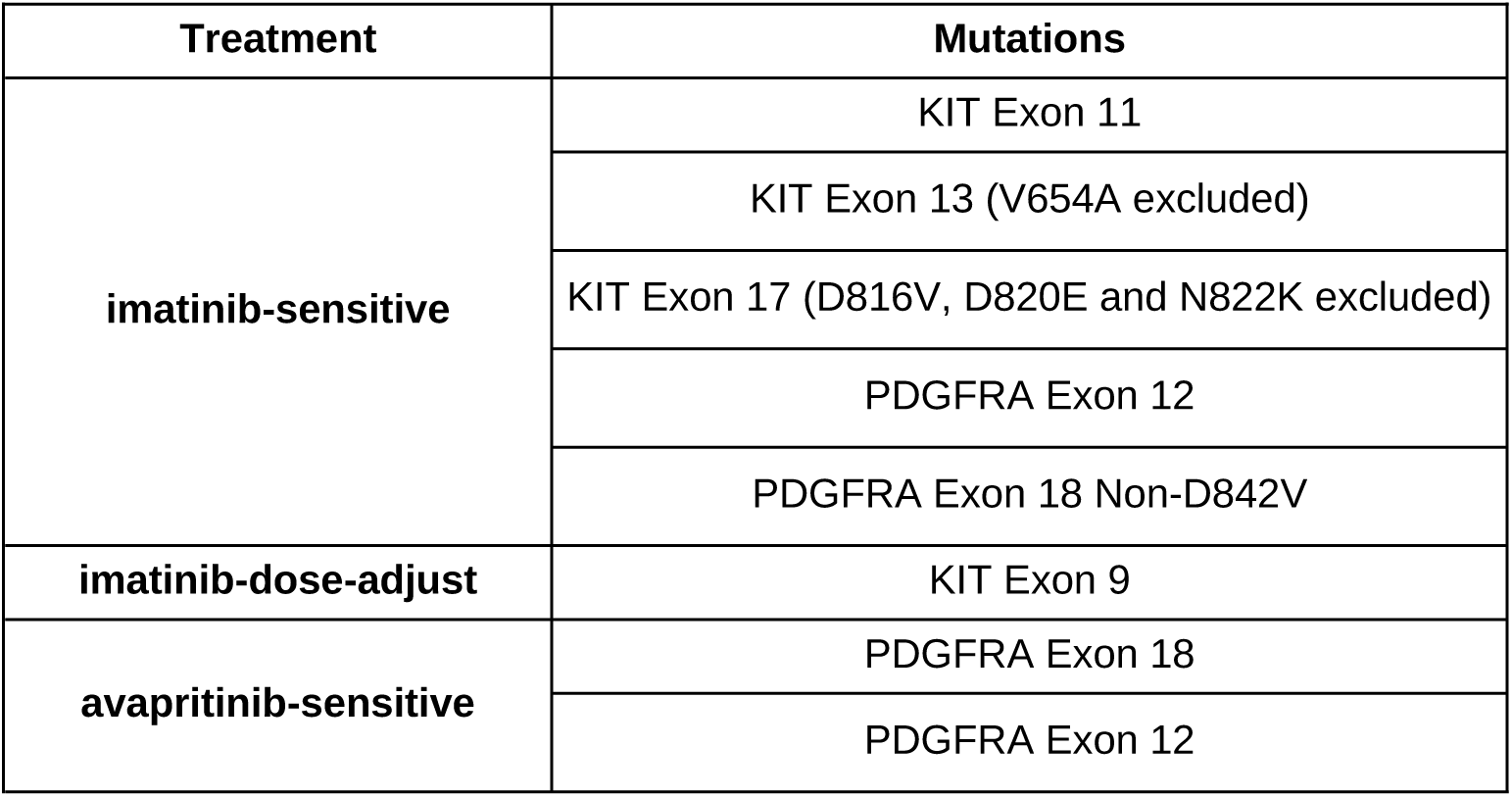
Grouping of mutations according to treatment sensitivity categories used for DL-based prediction. The table defines how specific *KIT* and *PDGFRA* mutations were classified as imatinib-sensitive, imatinib–dose-adjust, or avapritinib-sensitive

| Treatment | Mutations |
| --- | --- |
| imatinib-sensitive | KIT Exon 11 |
|  | KIT Exon 13 (V654A excluded) |
|  | KIT Exon 17 (D816V, D820E and N822K excluded) |
|  | PDGFRA Exon 12 |
|  | PDGFRA Exon 18 Non-D842V |
| imatinib-dose-adjust | KIT Exon 9 |
| avapritinib-sensitive | PDGFRA Exon 18 |
|  | PDGFRA Exon 12 |

**Supplementary Table 6.** Performance of the ML models for predicting treatment sensitivity categories in GIST. For each category, the table reports accuracy, precision, recall, F1 score, and the AUC with 95% CI.

| Treatment | Accuracy | Precision | Recall | F1 | AUC | CI_lower | CI_upper |
| --- | --- | --- | --- | --- | --- | --- | --- |
| imatinib-sensitive | 0.65 | 0.65 | 1.00 | 0.79 | 0.59 | 0.52 | 0.65 |
| imatinib-dose-adjust | 0.61 | 0.16 | 0.90 | 0.28 | 0.76 | 0.68 | 0.84 |
| avapritinib-sensitive | 0.08 | 0.08 | 1.00 | 0.15 | 0.54 | 0.45 | 0.65 |

**Supplementary Table 7.**
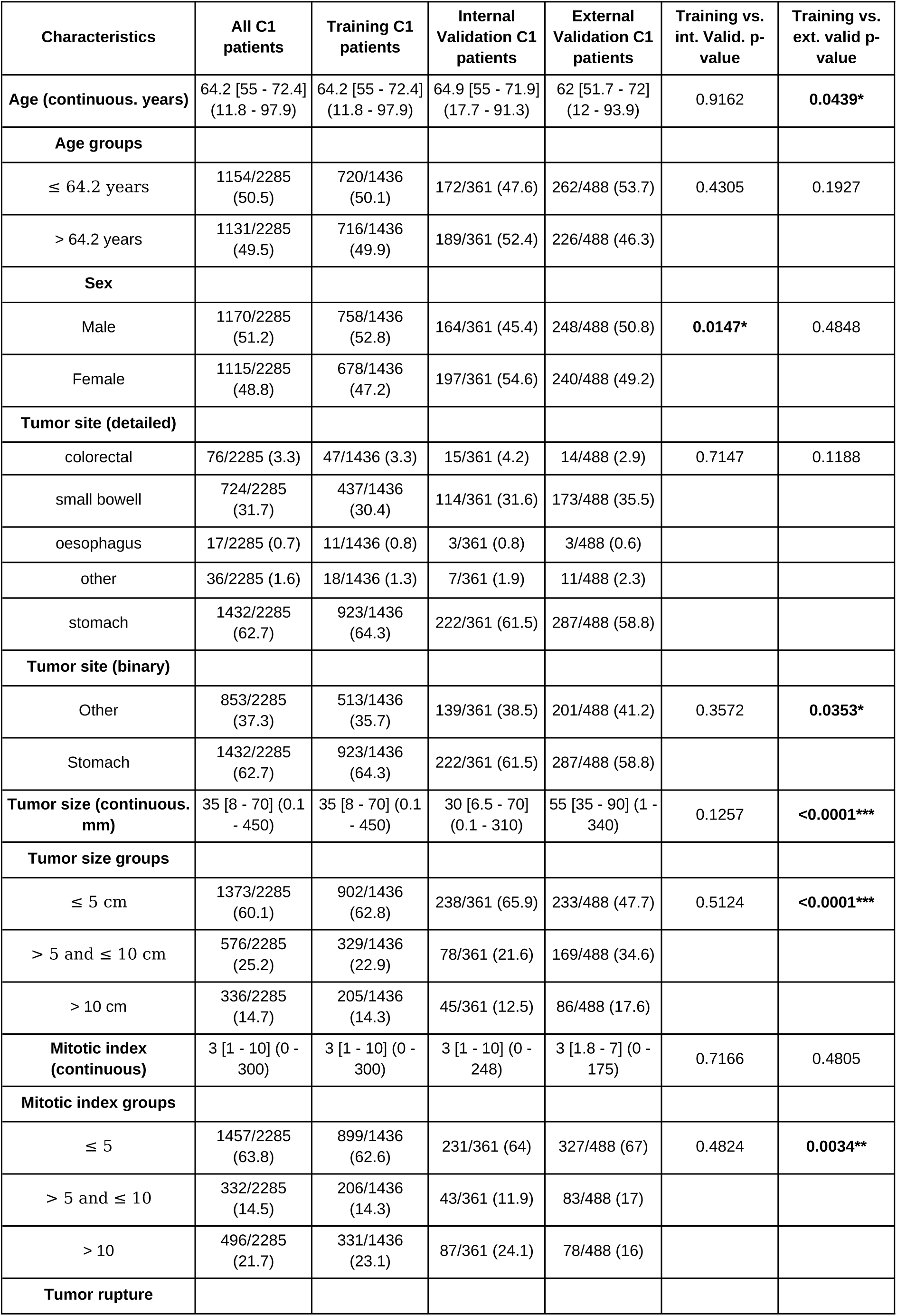
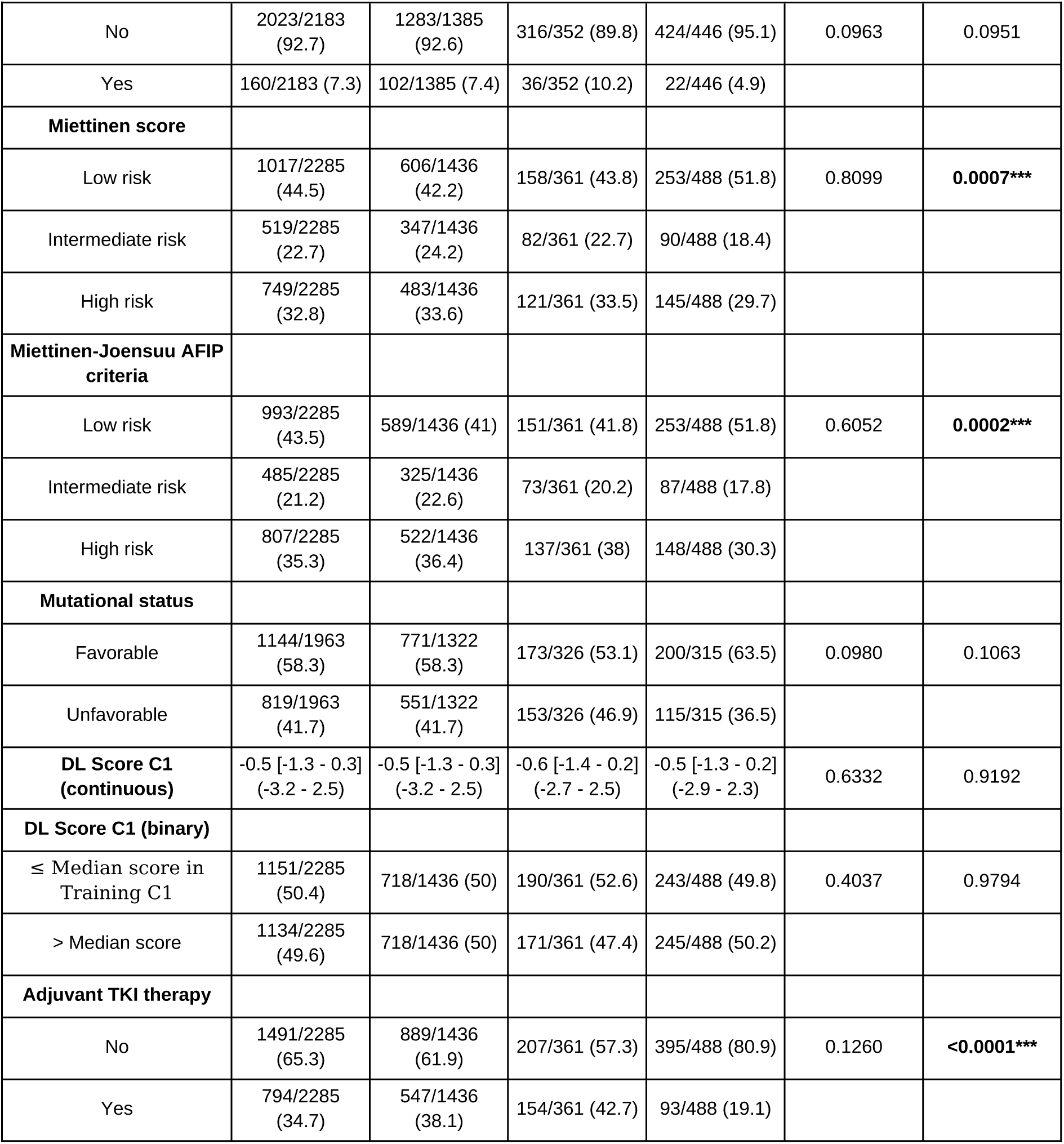
Baseline characteristics of patients from the C1 cohort included in the recurrence-free survival (RFS) analysis. Data are presented as numbers with percentages in parentheses for categorical variables and as medians with interquartile and minimum–maximum ranges for continuous variables. Comparisons of baseline characteristics were performed between the training and internal validation (int. val.) sets and between the training and external validation (ext. val.) sets using Chi-square or Mann–Whitney tests, as appropriate. *p < 0.05; **p < 0.005; ***p < 0.001.

**Supplementary Table 8.**
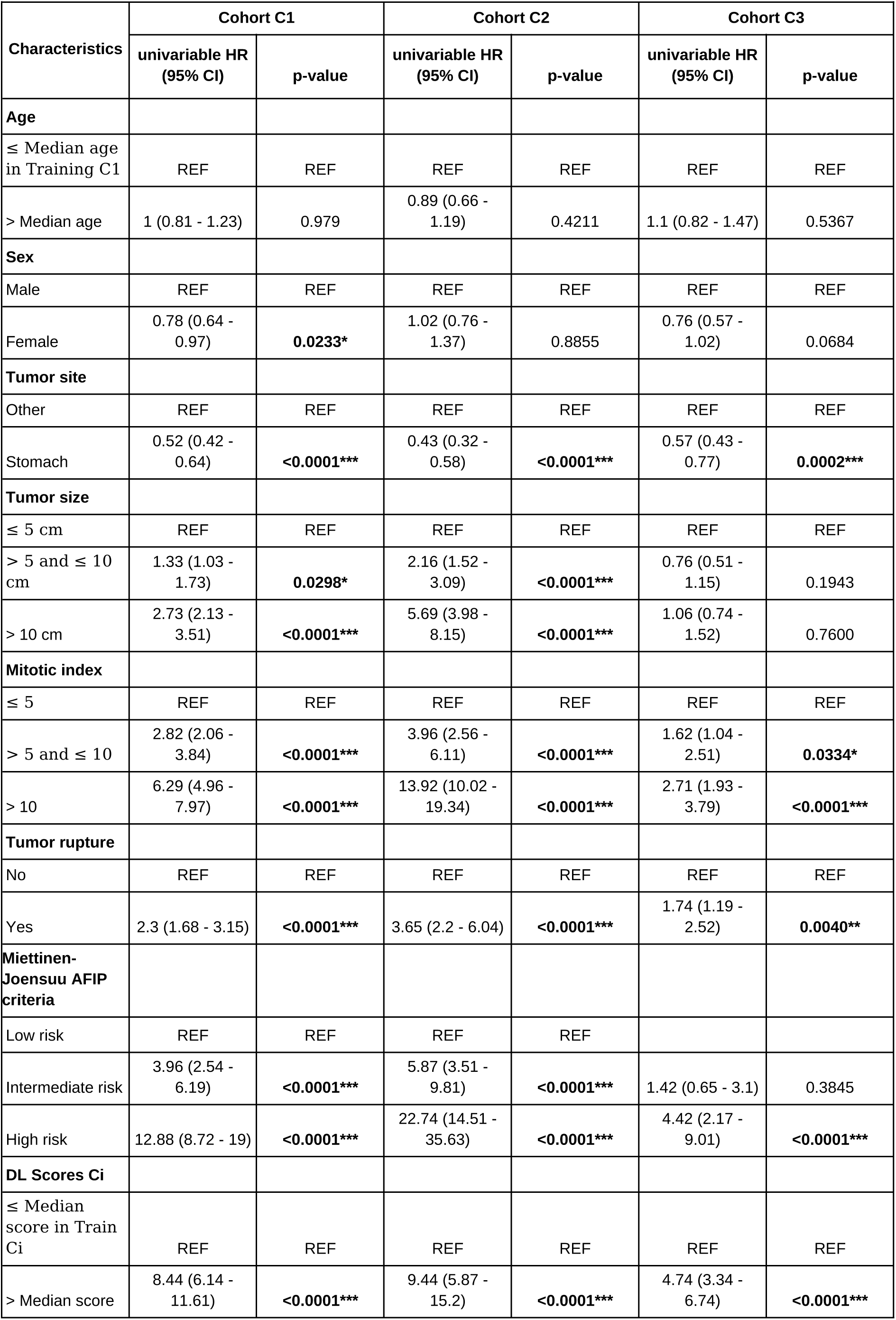
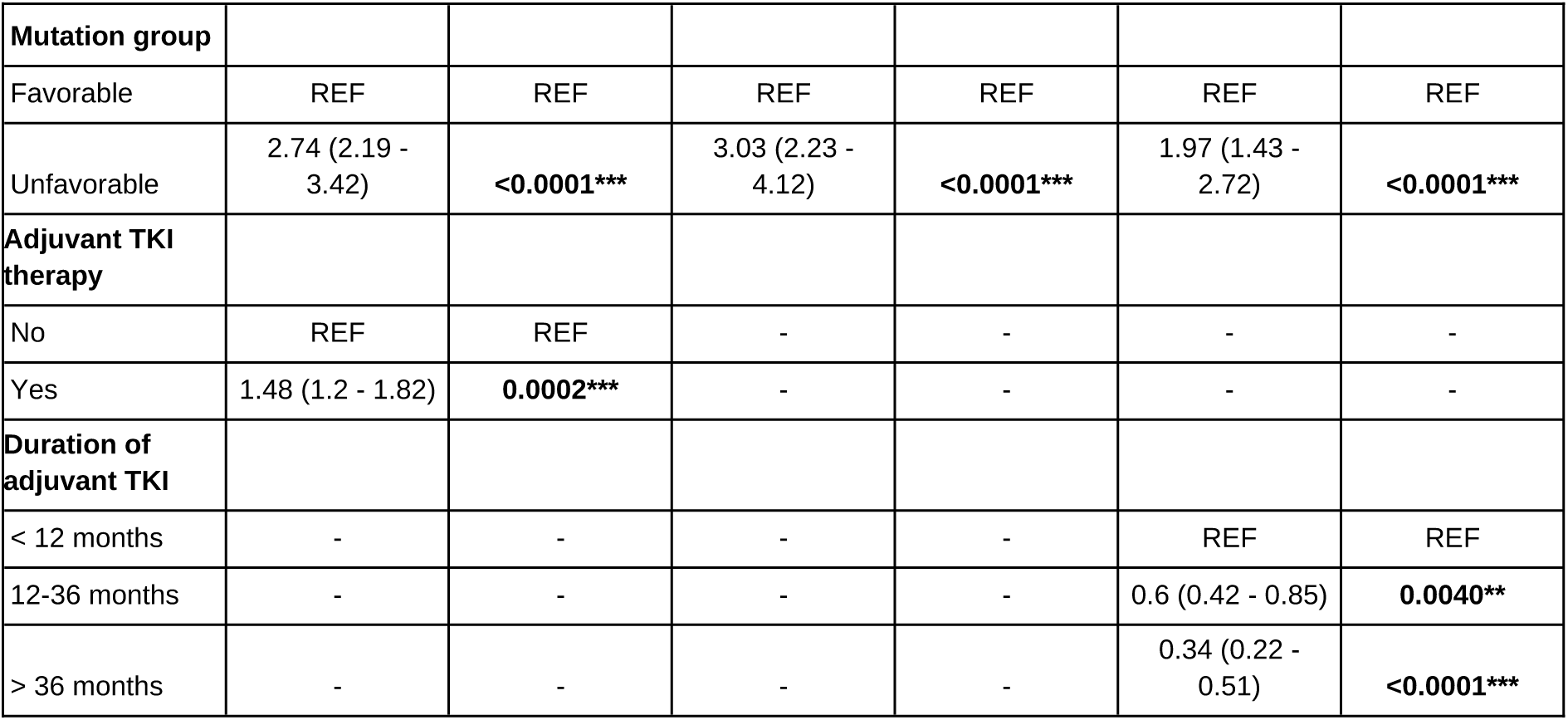
Univariable recurrence-free survival (RFS) analysis in the Training datasets from the C1, C2 and C3 cohorts. Other abbreviations: CI: confidence interval, DL: deep learning, REF: level of reference for univariable Cox regression, TKI: tyrosine kinase inhibitor. *: p < 0.05; **: p < 0.005; ***: p < 0.001.

**Supplementary Table 9.**
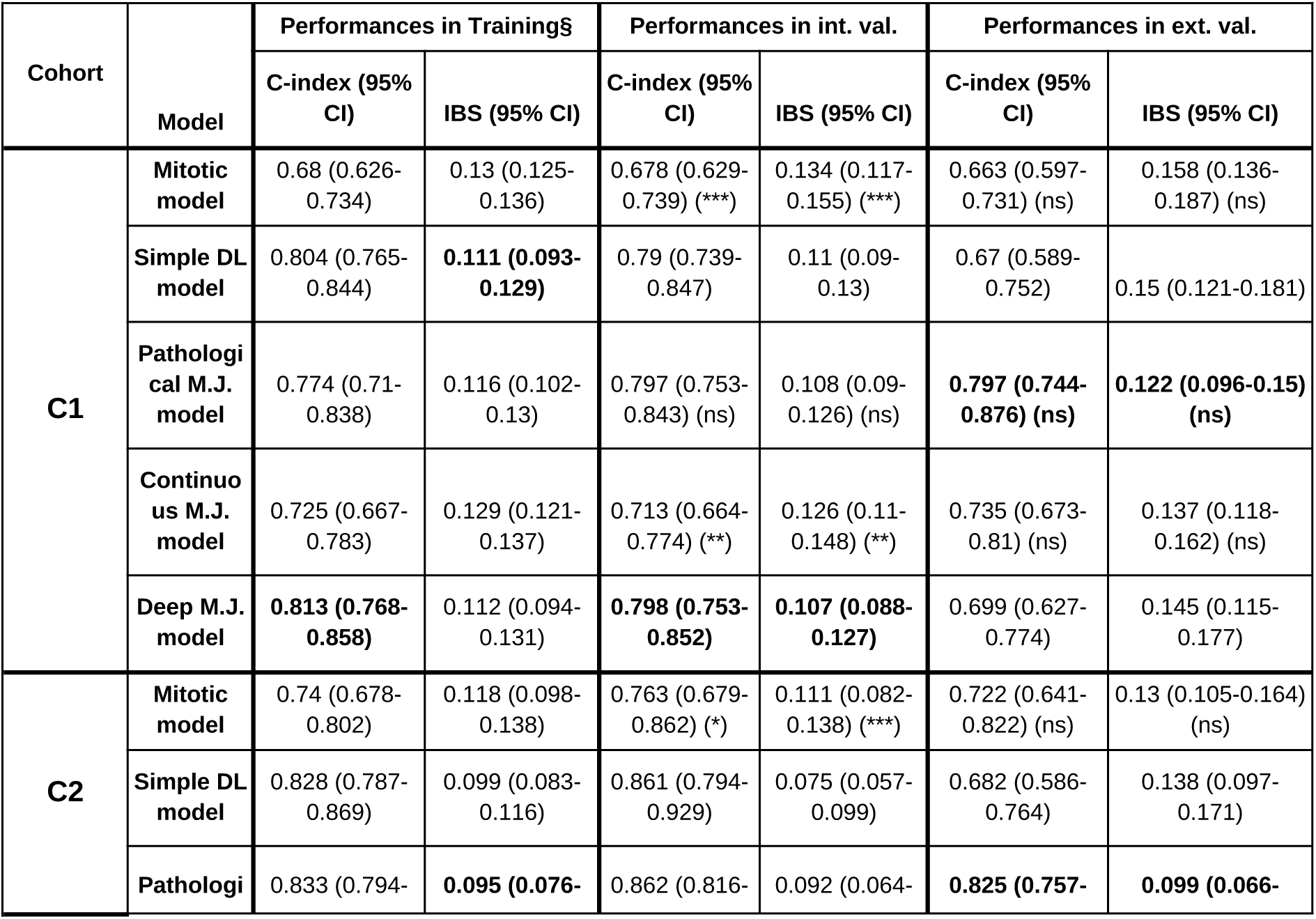
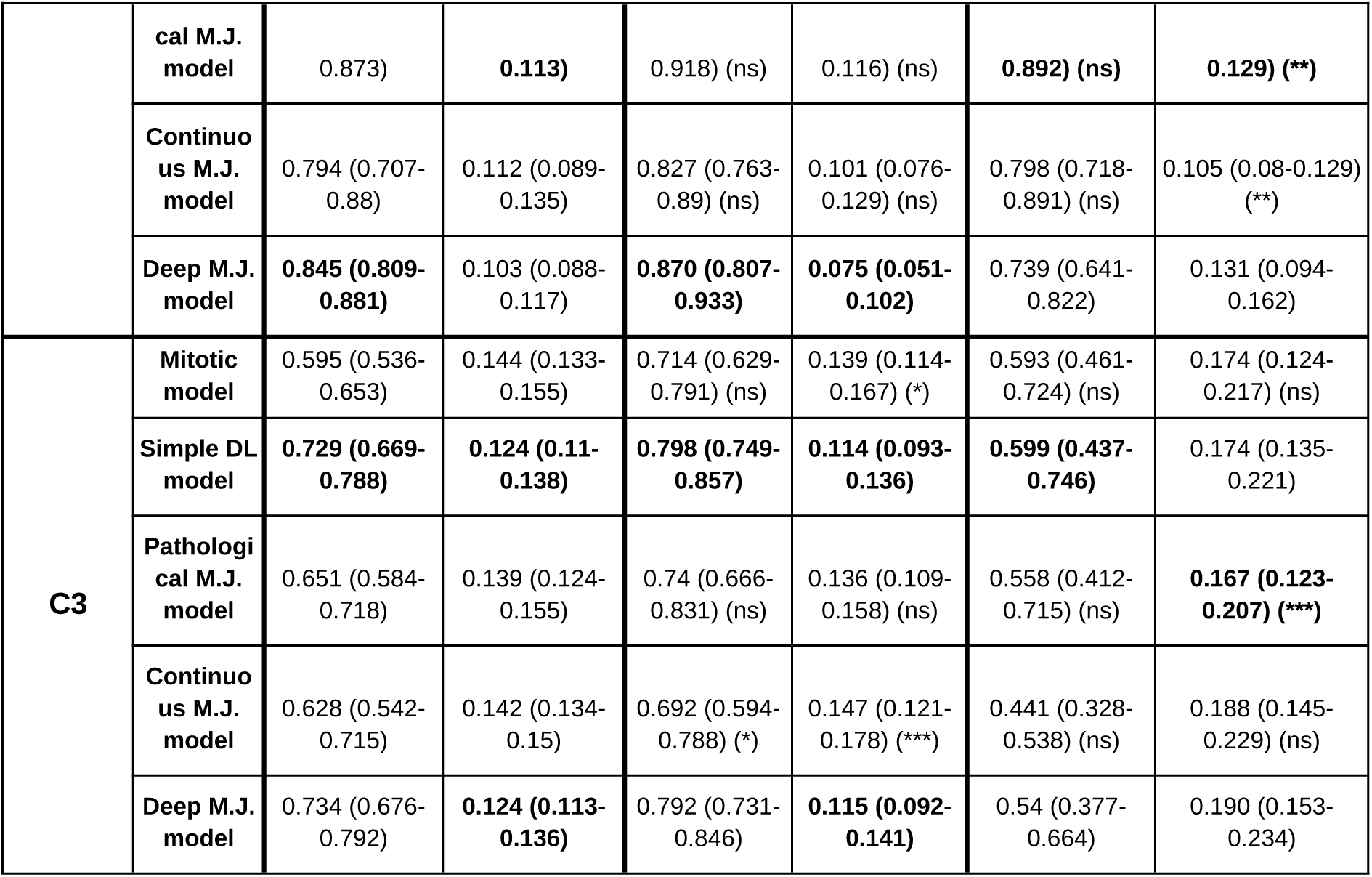
Prognostic performances of survival models across cohorts C1–C3. Concordance indices (C-index) and integrated Brier scores (IBS) with 95% confidence intervals (CIs) are reported for the mitotic index–based model (“Mitotic”), the simple deep learning (DL) score model (“Simple DL”), the pathological Miettinen–Joensuu model (“Path. MJ”), the continuous Miettinen–Joensuu model (“Cont. MJ”), and the deep Miettinen–Joensuu model (“Deep MJ”) in training (out-of-fold cross-validation), internal validation, and external validation datasets. Results are shown for the full C1 cohort, the C2 subcohort (patients without adjuvant TKI therapy), and the C3 subcohort (patients with adjuvant TKI therapy). Significance levels from permutation tests comparing paired models are indicated as follows: ns, not significant; *p < 0.05; **p < 0.005; ***p < 0.001.

**Supplementary Table 10.**
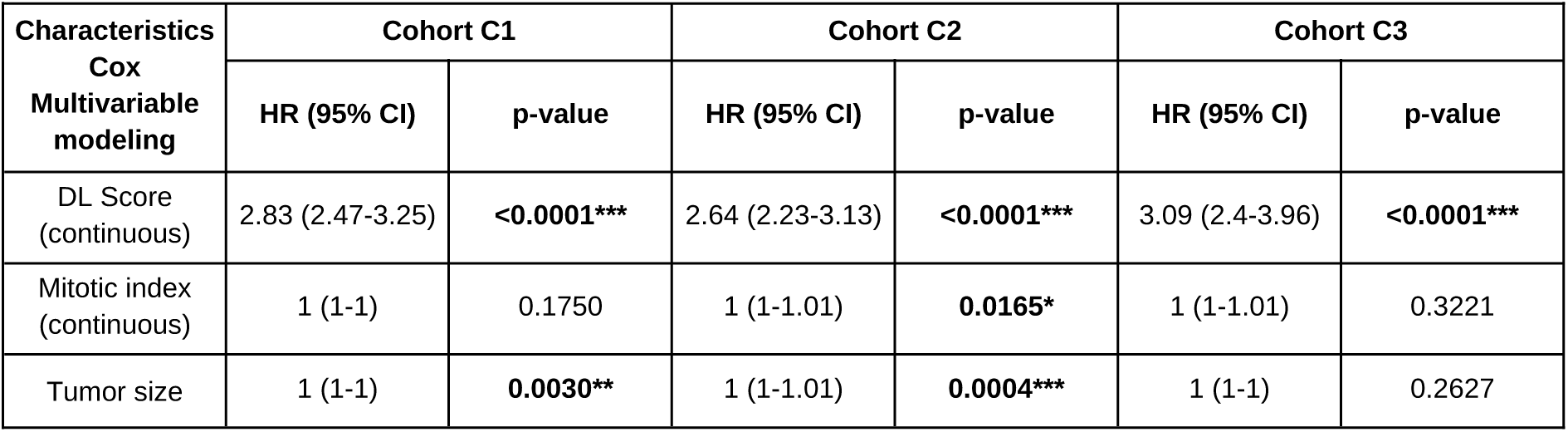
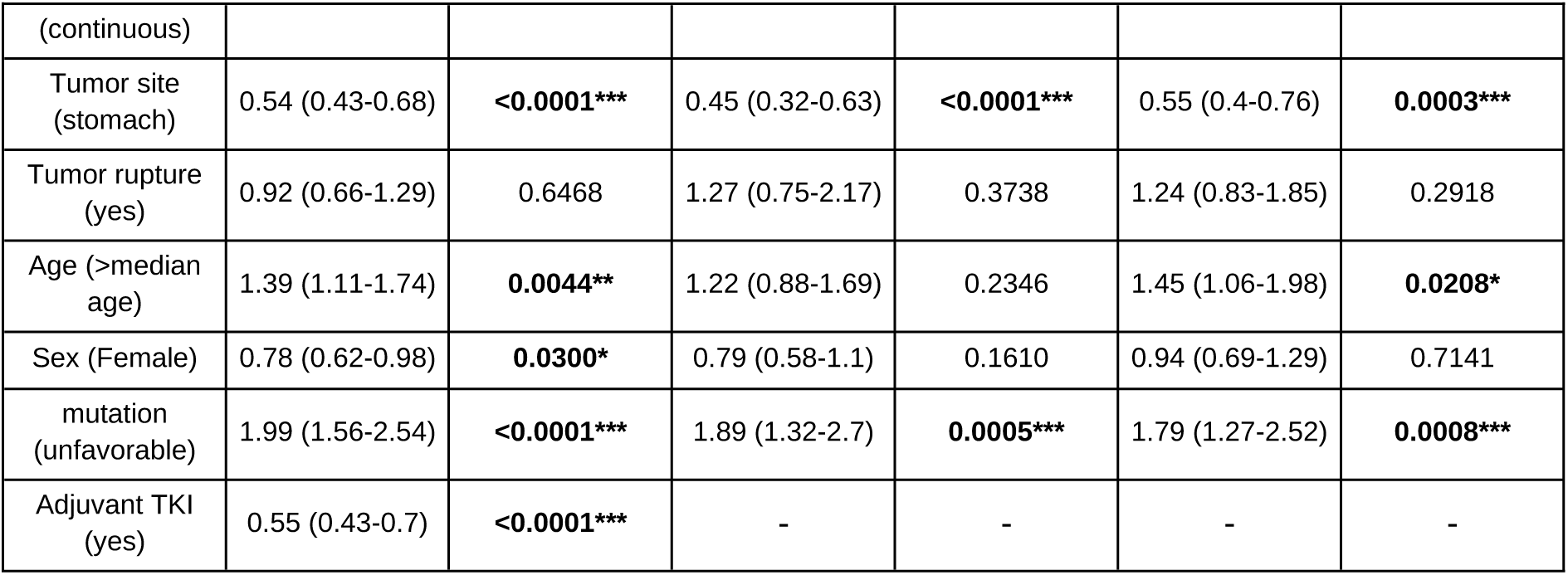
Multivariable Cox regression analyses in cohorts C1–C3. Hazard ratios (HRs) with 95% confidence intervals (CIs) and p-values are reported for the deep learning (DL) score (continuous), mitotic index (continuous), tumor size (continuous), tumor site (stomach vs. other), tumor rupture, age (>median), sex (female vs. male), mutation status (unfavorable vs. favorable), and adjuvant TKI therapy (yes vs. no). Results are shown for the overall C1 cohort, the C2 subcohort (patients without adjuvant TKI therapy), and the C3 subcohort (patients with adjuvant TKI therapy). The DL score remained an independent prognostic factor in all cohorts, with hazard ratios ranging from 2.64 to 3.09. Other abbreviations: TKI, tyrosine kinase inhibitor. *p < 0.05; **p < 0.005; ***p < 0.001. Significant results are in bold.

**Supplementary Table 11.**
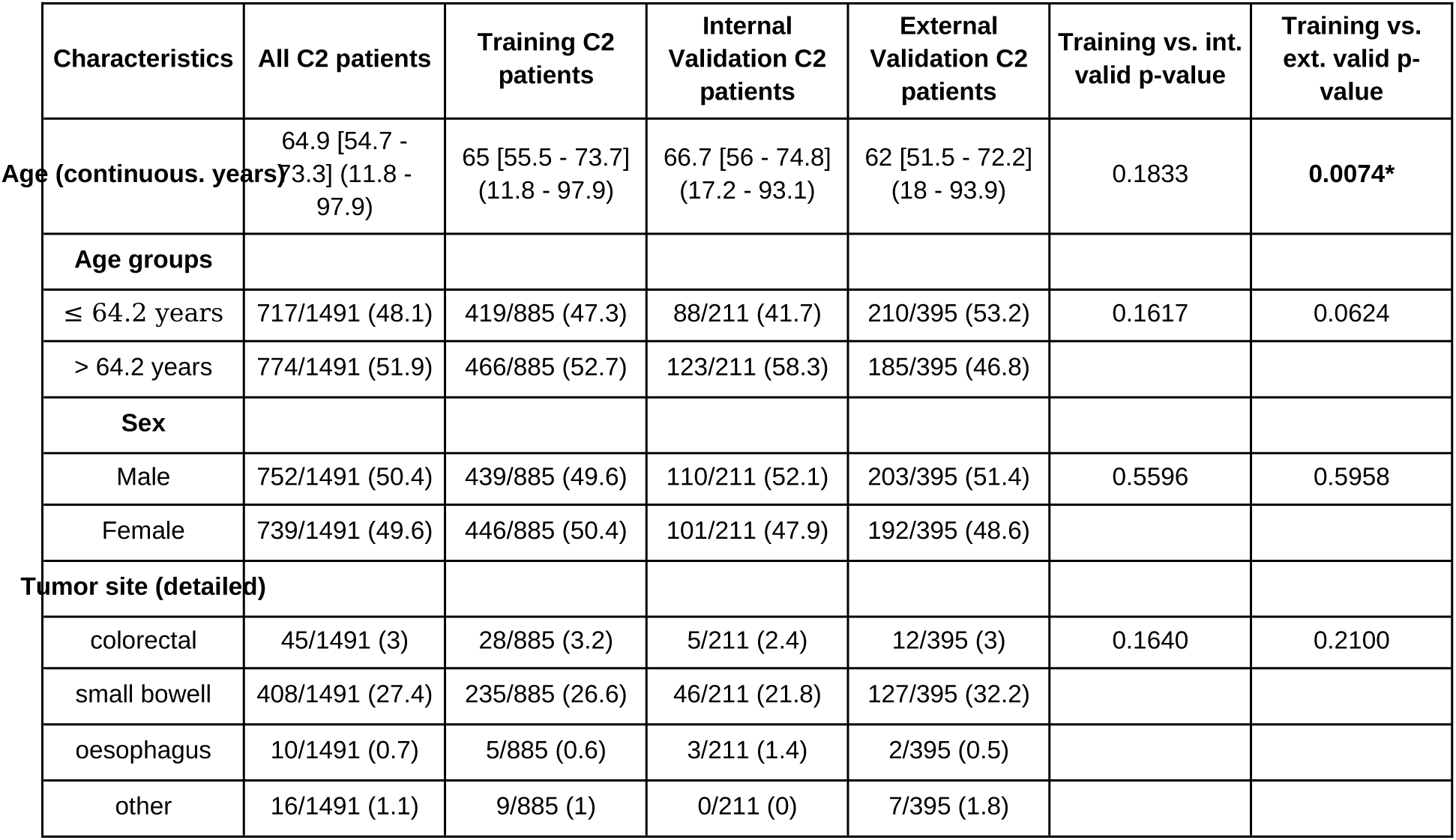
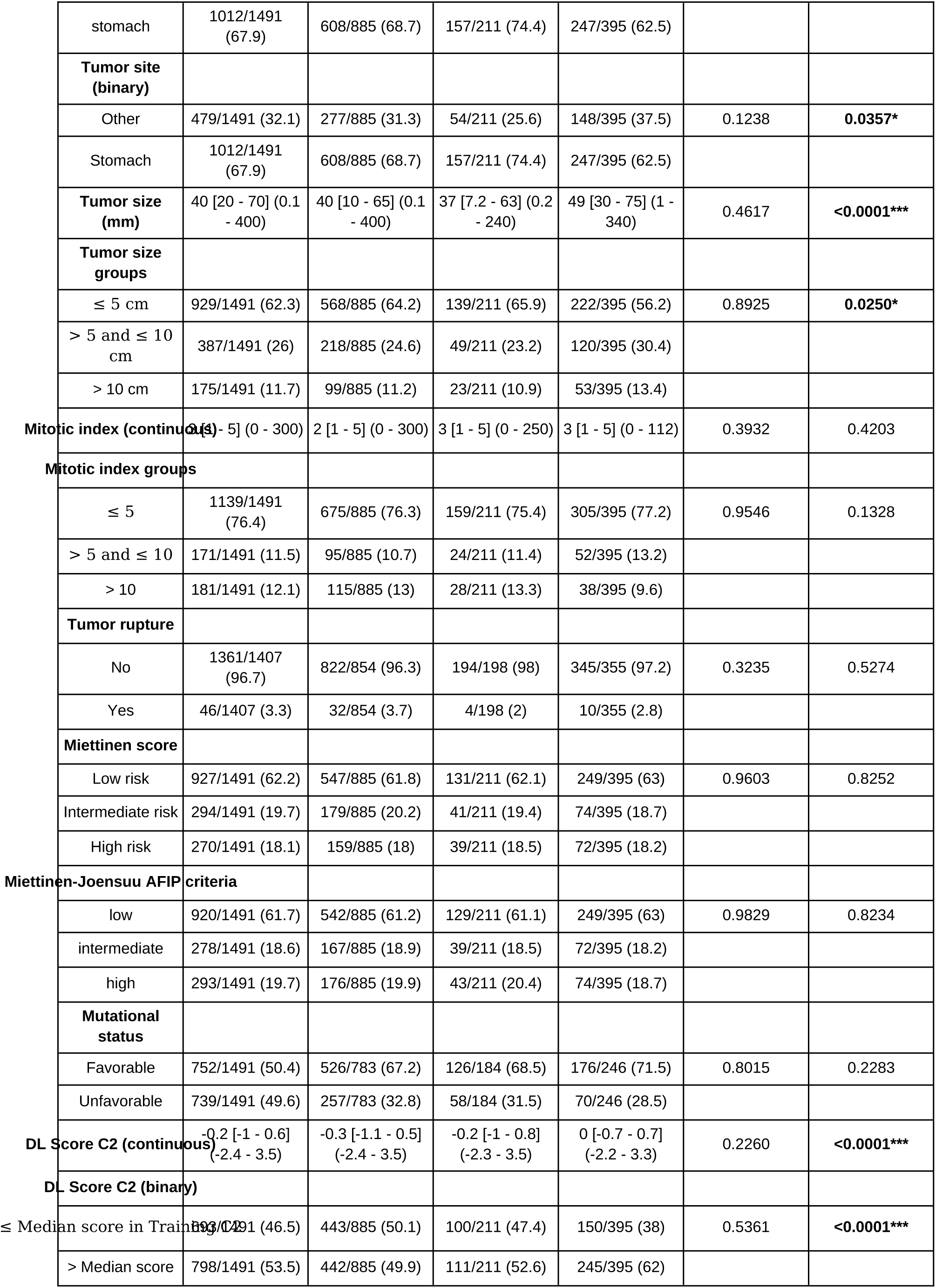
Baseline characteristics of patients from the C2 cohort included in the recurrence-free survival (RFS) analysis. Data are presented as numbers with percentages in parentheses for categorical variables and as medians with interquartile and minimum–maximum ranges for continuous variables. Comparisons of baseline characteristics were performed between the training and internal validation (int. val.) sets and between the training and external validation (ext. val.) sets using Chi-square or Mann–Whitney tests, as appropriate. *p < 0.05; **p < 0.005; ***p < 0.001.

**Supplementary Table 12.**
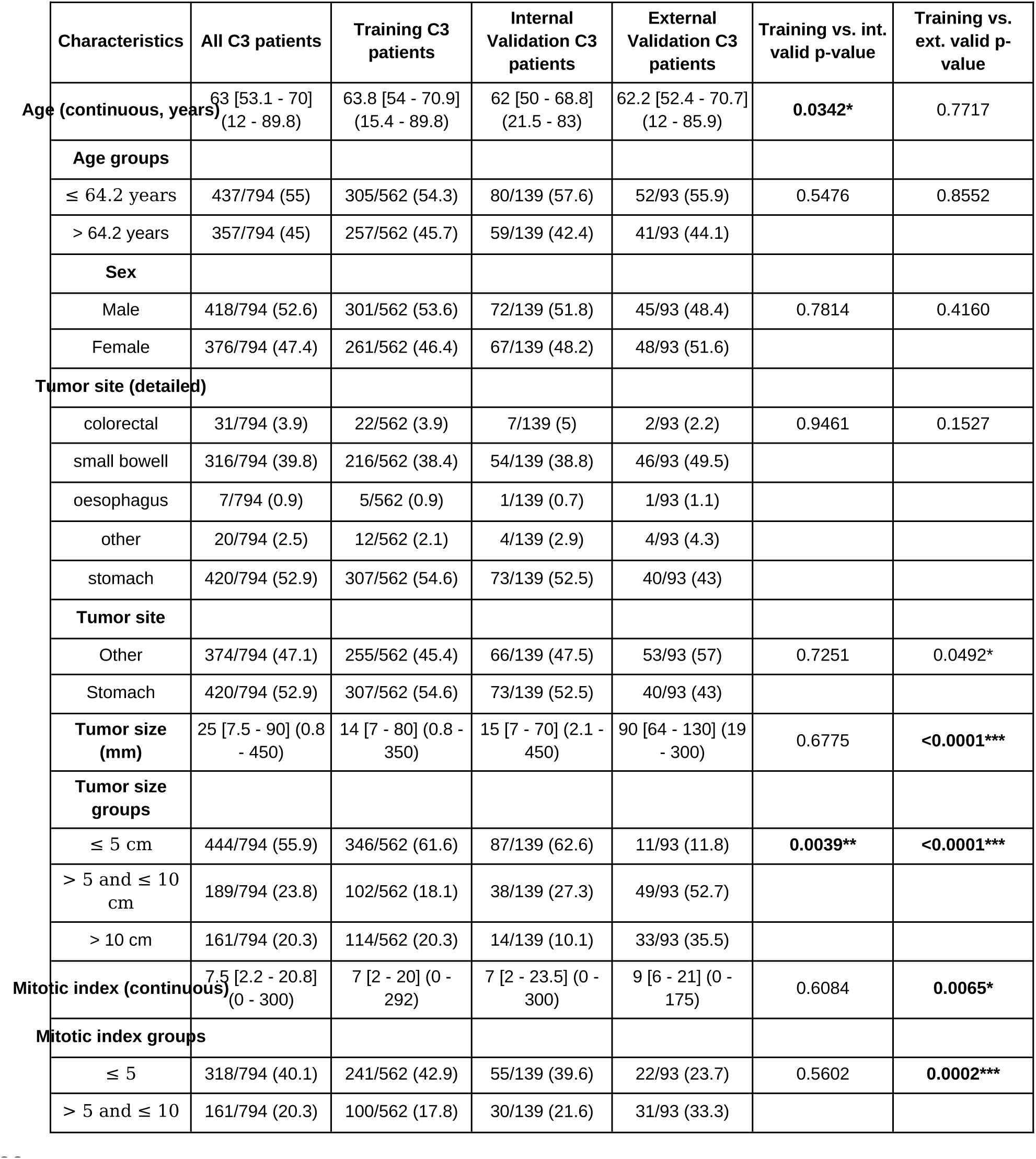
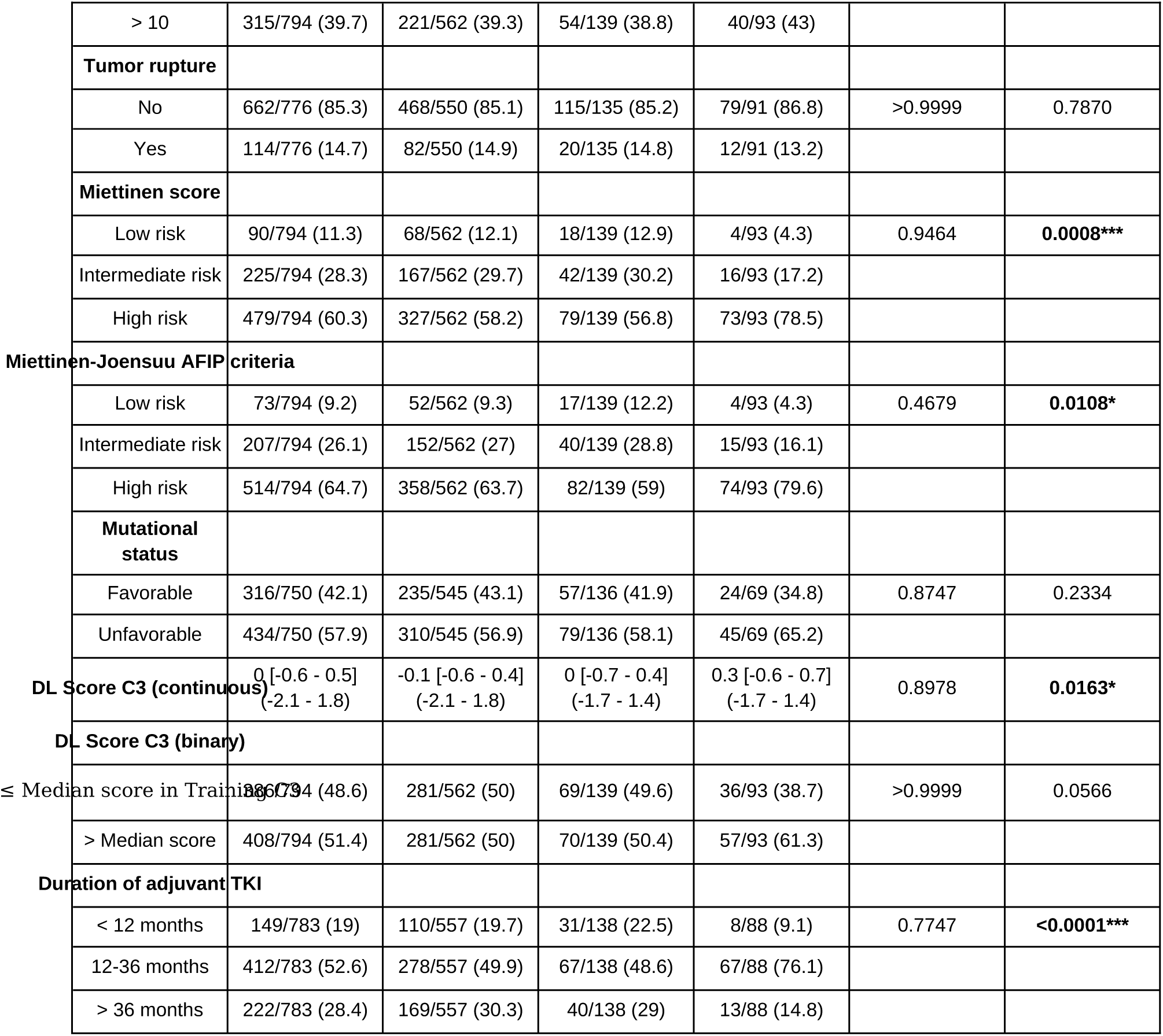
Baseline characteristics of patients from the C3 cohort included in the recurrence-free survival (RFS) analysis. Data are presented as numbers with percentages in parentheses for categorical variables and as medians with interquartile and minimum–maximum ranges for continuous variables. Comparisons of baseline characteristics were performed between the training and internal validation (int. val.) sets and between the training and external validation (ext. val.) sets using Chi-square or Mann–Whitney tests, as appropriate. *p < 0.05; **p < 0.005; ***p < 0.001.

**Supplementary Table 13.** Recurrence-free survival (RFS) and overall survival (OS) in patients from C2 and C3 classified as high risk according to the pathological Miettinen–Joensuu (MJ) scoring system and carrying imatinib-sensitive mutations. Hazard ratios (HRs) with 95% confidence intervals (CIs) are shown; REF indicates the reference category. *p < 0.05; **p < 0.005; ***p < 0.001. Significant results are in bold.

| Outcome | Characteristics |  | Survival probability at 5 years | log-rank p-value | HR (95% CI) | Cox p-value |
| --- | --- | --- | --- | --- | --- | --- |
| RFS | Deep MJ C2 groups |  |  |  |  |  |
|  |  | ≤ median | 43.77 (33.72 - 56.82) | **0.0007\*\*\*** | REF | REF |
|  |  | > median | 23.12 (15.42 - 34.66) |  | 1.86 (1.29 - 2.67) | **0.0008\*\*\*** |
|  | Deep MJ C3 groups |  |  |  |  |  |
|  |  | ≤ median | 78.96 (73.3 - 85.05) | **<0.0001\*\*\*** | REF | REF |
|  |  | > median | 51.03 (44.5 - 58.52) |  | 3.44 (2.5 - 4.72) | **<0.0001\*\*\*** |
|  | Summary deep MJ C2 and C3 groups |  |  |  |  |  |
|  |  | C2 - low risk<br>Deep MJ C2 | 43.77 (33.72 - 56.82) | **<0.0001\*\*\*** | REF | REF |
|  |  | C2 - high risk<br>Deep MJ C2 | 23.12 (15.42 - 34.66) |  | 0.6 (0.47 - 0.75) | **<0.0001\*\*\*** |
|  |  | C3 - low_risk<br>Deep MJ C3 | 78.96 (73.3 - 85.05) |  | 1.29 (1.02 - 1.65) | **0.0352\*** |
|  |  | C3 - high risk<br>Deep MJ C3 | 51.03 (44.5 - 58.52) |  | 3.59 (2.8 - 4.59) | **<0.0001\*\*\*** |
| OS | Deep MJ C2 groups |  |  |  |  |  |
|  |  | ≤ median | 88.69 (81.46 - 96.56) | **0.0333\*** | REF | REF |
|  |  | > median | 76.94 (68.07 - 86.95) |  | 1.78 (1.04 - 3.06) | **0.0356\*** |
|  | Deep MJ C3 groups |  |  |  |  |  |
|  |  | ≤ median | 95.13 (92.22 - 98.12) | **<0.0001\*\*\*** | REF | REF |
|  |  | > median | 84.78 (79.87 - 89.99) |  | 3.44 (2.04 - 5.79) | **<0.0001\*\*\*** |
|  | Summary deep MJ C2 and C3 groups |  |  |  |  |  |
|  |  | C2 - low risk<br>Deep MJ C2 | 88.69 (81.46 - 96.56) | **<0.0001\*\*\*** | REF | REF |
|  |  | C2 - high risk<br>Deep MJ C2 | 76.94 (68.07 - 86.95) |  | 0.85 (0.59 - 1.23) | 0.3987 |
|  |  | C3 - low_risk<br>Deep MJ C3 | 95.13 (92.22 - 98.12) |  | 1.39 (0.96 - 2.02) | 0.085 |
|  |  | C3 - high risk<br>Deep MJ C3 | 84.78 (79.87 - 89.99) |  | 2.99 (2.03 - 4.39) | **<0.0001\*\*\*** |

## References

1. International Agency for Research on Cancer, World Health Organization, International Academy of Pathology. WHO classification of tumours of soft tissue and bone tumours, 5th edition. Fletcher CDM (ed): IARC 2020. (World Health Organization Classification of Tumours).

2. Casali PG, Blay JY, Abecassis N, Bajpai J, Bauer S, Biagini R, et al. Gastrointestinal stromal tumours: ESMO-EURACAN-GENTURIS Clinical Practice Guidelines for diagnosis, treatment and follow-up. Ann Oncol. January 1, 2022;33(1):20–33.

3. Søreide K, Sandvik OM, Søreide JA, Giljaca V, Jureckova A, Bulusu VR. Global epidemiology of gastrointestinal stromal tumours (GIST): A systematic review of population-based cohort studies. Cancer Epidemiol. February 1, 2016;40:39–46.

4. Gastrointestinal Stromal Tumors Treatment (PDQ®) [Internet]. 2011 [cited February 24, 2025]. Available at: https://www.cancer.gov/types/soft-tissue-sarcoma/hp/gist-treatment-pdq

5. American Joint Committee on Cancer. AJCC cancer staging manual, 7th edition. London, England: Springer 2010.

6. Hirota S, Isozaki K, Moriyama Y, Hashimoto K, Nishida T, Ishiguro S, et al. Gain-of-function mutations of c-kit in human gastrointestinal stromal tumors. Science. January 23, 1998;279(5350):577–580.

7. Corless CL, Barnett CM, Heinrich MC. Gastrointestinal stromal tumours: origin and molecular oncology. Nat Rev Cancer. November 17, 2011;11(12):865–878.

8. Blay J-Y, Kang Y-K, Nishida T, von Mehren M. Gastrointestinal stromal tumours. Nat Rev Dis Primers. March 18, 2021;7(1):22.

9. Penel N, Le Cesne A, Blay J-Y. Adjuvant treatment of gastrointestinal stromal tumor: State of the art in 2025. Eur J Cancer. June 3, 2025;222(115473):115473.

10. Tsai H-J, Shan Y-S, Yang C-Y, Hsiao C-F, Tsai C-H, Wang C-C, et al. Survival of advanced/recurrent gastrointestinal stromal tumors treated with tyrosine kinase inhibitors in Taiwan: a nationwide registry study. BMC Cancer. July 11, 2024;24(1):828.

11. Fu Y, Karanian M, Perret R, Camara A, Le Loarer F, Jean-Denis M, et al. Deep learning predicts patients outcome and mutations from digitized histology slides in gastrointestinal stromal tumor. NPJ Precis Oncol. July 24, 2023;7(1):71.

12. Kong X, Shi J, Sun D, Cheng L, Wu C, Jiang Z, et al. A deep-learning model for predicting tyrosine kinase inhibitor response from histology in gastrointestinal stromal tumor. J Pathol. April 2025;265(4):462–471.

13. Judson I, Jones RL, Wong NACS, Dileo P, Bulusu R, Smith M, et al. Gastrointestinal stromal tumour (GIST): British Sarcoma Group clinical practice guidelines. Br J Cancer. January 2025;132(1):1–10.

14. Mavroeidis L, Kalofonou F, Casey R, Napolitano A, Bulusu R, Jones RL. Identifying and managing rare subtypes of gastrointestinal stromal tumors. Expert Rev Gastroenterol Hepatol. April 2025;19(5):549–561.

15. Joensuu H, Vehtari A, Riihimäki J, Nishida T, Steigen SE, Brabec P, et al. Risk of recurrence of gastrointestinal stromal tumour after surgery: an analysis of pooled population-based cohorts. Lancet Oncol. March 2012;13(3):265–274.

16. Bertsimas D, Margonis GA, Tang S, Koulouras A, Antonescu CR, Brennan MF, et al. An interpretable AI model for recurrence prediction after surgery in gastrointestinal stromal tumour: an observational cohort study. EClinicalMedicine. October 2023;64(102200):102200.

17. Liang H, Li Z, Lin W, Xie Y, Zhang S, Li Z, et al. Enhancing gastrointestinal stromal tumor (GIST) diagnosis: An improved YOLOv8 deep learning approach for precise mitotic detection. IEEE Access. 2024;12:116829–116840.

18. Brink P, Kalisvaart GM, Schrage YM, Mohammadi M, Ijzerman NS, Bleckman RF, et al. Local treatment in metastatic GIST patients: A multicentre analysis from the Dutch GIST Registry. Eur J Surg Oncol. September 2023;49(9):106942.

19. Bertsimas D, Margonis GA, Sujichantararat S, Koulouras A, Ma Y, Antonescu CR, et al. Interpretable artificial intelligence to optimise use of imatinib after resection in patients with localised gastrointestinal stromal tumours: an observational cohort study. Lancet Oncol. August 2024;25(8):1025–1037.

20. Zeng Q, Klein C, Caruso S, Maille P, Laleh NG, Sommacale D, et al. Artificial intelligence predicts immune and inflammatory gene signatures directly from hepatocellular carcinoma histology. J Hepatol. July 2022;77(1):116–127.

21. El Nahhas OSM, van Treeck M, Wölflein G, Unger M, Ligero M, Lenz T, et al. From whole-slide image to biomarker prediction: end-to-end weakly supervised deep learning in computational pathology. Nat Protoc. January 2025;20(1):293–316.

22. Lu MY, Chen B, Williamson DFK, Chen RJ, Liang I, Ding T, et al. A visual-language foundation model for computational pathology. Nat Med. March 19, 2024;30(3):863–874.

23. Dosovitskiy A, Beyer L, Kolesnikov A, Weissenborn D, Zhai X, Unterthiner T, et al. An image is worth 16×16 words: Transformers for image recognition at scale [Internet]. arXiv [cs.CV]. 2020. Available at: http://arxiv.org/abs/2010.11929

24. Wang M, Callenberg KM, Dalgleish R, Fedtsov A, Fox NK, Freeman PJ, et al. hgvs: A Python package for manipulating sequence variants using HGVS nomenclature: 2018 Update. Hum Mutat. December 1, 2018;39(12):1803–1813.

25. Lenz T, Neidlinger P, Ligero M, Wölflein G, van Treeck M, Kather JN. Unsupervised foundation model-agnostic slide-level representation learning [Internet]. arXiv [cs.CV]. 2024. Available at: http://arxiv.org/abs/2411.13623

26. Selvaraju RR, Cogswell M, Das A, Vedantam R, Parikh D, Batra D. Grad-CAM: Visual explanations from deep networks via Gradient-based localization [Internet]. arXiv [cs.CV]. 2016. Available at: http://arxiv.org/abs/1610.02391

27. Blay J-Y, Schiffler C, Bouché O, Brahmi M, Duffaud F, Toulmonde M, et al. A randomized study of 6 versus 3 years of adjuvant imatinib in patients with localized GIST at high risk of relapse. Ann Oncol. December 2024;35(12):1157–1168.

28. Briercheck EL, Wrigglesworth JM, Garcia-Gonzalez I, Scheepers C, Ong MC, Venkatesh V, et al. Treatment access for gastrointestinal stromal tumor in predominantly low– and middle-income countries. JAMA Netw Open. April 1, 2024;7(4):e244898.

29. Joensuu H, Rutkowski P, Nishida T, Steigen SE, Brabec P, Plank L, et al. KIT and PDGFRA mutations and the risk of GI stromal tumor recurrence. J Clin Oncol. February 20, 2015;33(6):634–642.

